# Geospatial assessment of Japanese encephalitis virus infection risk in Australia

**DOI:** 10.64898/2026.10.15.26365607

**Authors:** Lucinda E. Harrison, David H. Duncan, Jennifer Flegg, David J. Price, Nina Kurucz, Ximena Tolosa, Peter J. Neville, Cassie C. Jansen, Abbey Potter, Craig Brockway, Kimberly L. Miller, Michaela Hobby, Vicki Burns, Keira M. Glasgow, Anthony D. K. Draper, Josef Schmidt, Guy Weerasinghe, Andrew F. van den Hurk, David T. Williams, Stephen L. Doggett, Nick Golding, Freya M. Shearer

**Affiliations:** School of Mathematics and Statistics, University of Melbourne, Parkville, VIC, Australia; Infectious Disease Dynamics Unit, Centre for Epidemiology and Biostatistics, Melbourne School of Population and Global Health, University of Melbourne, Parkville, VIC, Australia; Infectious Diseases Data Observatory, Centre for Global Health Research, Nuffield Department of Medicine, University of Oxford, Oxford, United Kingdom; Infectious Disease Ecology and Modelling, The Kids Research Institute Australia, Nedlands, WA, Australia; Peter Doherty Institute, Melbourne, Australia; Medical Entomology - Centre for Disease Control, Public Health Division, NT Health, Casuarina, NT, Australia; Public Health Intelligence Branch, Population Health Division, Queensland Department of Health, Brisbane, QLD, Australia; Vector-borne, Zoonotic and Environmental Health Diseases, Chief Health Officer Group, Department of Health, Melbourne, VIC, Australia; Health Protection and Regulation Branch, Queensland Department of Health, Brisbane, QLD, Australia; Environmental Health Directorate, Public and Aboriginal Health Division, Western Australia Department of Health, East Perth, WA, Australia; Health Protection and Regulation Branch, Department for Health and Wellbeing, SA Health, SA, Australia; Health Protection NSW, NSW Health, St Leonards, NSW, Australia; Centre for Disease Control, Northern Territory Government Department of Health, Darwin, NT, Australia; National Centre for Epidemiology and Population Health, College of Law, Governance and Policy, Australian National University, Canberra, ACT, Australia; Global and Tropical Health Division, Menzies School of Health Research, Charles Darwin University, Bidau Lecidere, Dili, Timor-Leste; Northern Australia Quarantine Strategy, Department of Agriculture, Fisheries and Forestry, Darwin, NT, Australia; Public Health Virology, Public and Environmental Health Reference Laboratories, Queensland Health, Coopers Plains, QLD, Australia; CSIRO, Australian Centre for Disease Preparedness, Geelong, VIC, Australia; Department of Medical Entomology, NSW Health Pathology, Westmead Hospital, Westmead, New South Wales, Australia; Physics, Maths and Computing, University of Western Australia, Crawley, WA, Australia

**Keywords:** Japanese encephalitis virus, arbovirus surveillance, emerging disease, geospatial modelling, OneHealth

## Abstract

Japanese encephalitis virus (JEV) is a leading cause of viral encephalitis in humans in Asia. Prior to 2021, only sporadic outbreaks had been detected in Australia’s tropical northern extremes. However, during 2021–23, JEV infection was identified in 45 people and in more than 85 piggeries, together with many detections in feral pigs, mosquitoes, and other livestock across Australia, including in the temperate far south. Here we estimated the spatial extent of virus transmission during the outbreak (the realised niche) and the area environmentally suitable for transmission (the potential niche). We collated a national dataset of JEV surveillance and detection in humans, mosquitoes, domestic pigs, feral pigs, and sentinel chickens from 2021–23. We combined these data with (1) previously published predictions of habitat suitability for vertebrate host and vector species, and (2) estimated densities of humans, domestic pigs, and feral pigs, in a multipart assessment of spatial risk. Our model projected broad potential environmental suitability over much of northern and eastern Australia during the outbreak period, but floodplain wetland areas in the north and the inland east of Australia had both the highest estimated environmental suitability for transmission, and the strongest evidence for virus circulation.

## Introduction

Japanese encephalitis virus (JEV) is a leading cause of viral encephalitis in humans in Asia. Globally, the disease is estimated to cause more than 20,000 deaths each year [1]; the symptomatic case fatality rate was recently estimated at between 11–17% [2]. Of symptomatic cases who survive the initial infection, 30–50 % suffer life-long neurological, cognitive, or behavioural complications [3]. The disease is zoonotic, with a number of confirmed and putative non-human vertebrate host species, including pigs and ardeid birds while humans are considered dead-end hosts. The virus is transmitted from host to host via a mosquito vector [4–6]. In Asia, Culex tritaeniorhynchus is a significant vector species, although evidence of vector competence and natural infection exists for a diverse group of mosquito species, particularly those belonging to the Culex genus [7]. The western extent of JEV transmission includes India and southern Pakistan, its northern extent includes eastern China and South Korea and, until 2021, its endemic range was considered to extend as far south as Indonesia and Papua New Guinea [8, 9]. Prior to 2021, JEV was only identified in Australia in the Torres Strait and Far North Queensland in sporadic outbreaks [5, 10], although surveillance since 2005 has been inconsistent [11]. Following a human case in the Tiwi Islands (off the coast of Darwin, Northern Territory) in February 2021, JEV spread widely across Australia during 2021–22 [12, 13]. Detections were recorded as far south as central Victoria, a temperate region more than 3,000 km from the Tiwi Islands. Genomic evidence suggests that the 2021–2023 outbreak resulted from a single reintroduction of JEV _independent_ of historical outbreaks [14, 15]. On the 4th of March 2022, the Acting Chief Medical Officer of Australia declared the outbreak of JEV in mainland Australia to be a Communicable Disease Incident of National Significance [16]. At that time, there was evidence of JEV exposure in 14 piggeries across the states of Queensland, New South Wales, Victoria and South Australia [16]. As of October 2023, there had been 45 human cases of JEV infection [17] (associated with 77 possible exposure sites), and evidence of JEV exposure in 123 piggeries, 20 pooled mosquito trap collections, at least 202 feral pigs, and 8 sentinel chicken flocks, together with evidence of exposure in horses and other livestock. Evidence of JEV transmission was recorded in the Northern Territory, Queensland, New South Wales, Victoria, South Australia, and the Kimberley and Pilbara regions of Western Australia; this area covers eastern, central, and north-western mainland Australia.

Understanding the spatial variation in disease risk is important for allocating human and animal health resources, including disease surveillance and control efforts. There are several ways to prevent or minimise the risk of JEV infection in humans, including the provision of vaccines to susceptible populations [11, 18], focal mosquito control [19], and public health advice encouraging personal protective measures to avoid mosquito bites [20, 21]. The size of the geographic area affected during the Australian 2021–23 outbreak and complexity associated with the timing and methods of surveillance make complete, systematic surveillance of the affected area practically impossible. Geospatial modelling is a useful tool to estimate spatial variation in underlying transmission risk by statistically accounting for potential spatial bias in detections [22–24]. Surveillance resources may then be geographically prioritised according to predicted risk (e.g., [25, 26]).

Another challenge for understanding spatial risk of JEV is uncertainty around the relative importance of different host and vector species to the JEV transmission cycle, and how this varies with mosquito species’ relative abundance, climate, and agricultural activities [27–29]. This uncertainty is particularly high in the Australian context, due to the virus’ recent emergence and the resulting absence of evidence for the involvement of putative host and vector species [5, 11, 30]. It is thought that the relative importance of different host and vector species in JEV transmission likely varies across the areas where JEV has been detected in Australia, which span biogeographic zones and include a range of habitats.

A number of works assessing spatial risk of JEV infection in Australia have been published in response to the 2021–23 outbreak [13, 31–35]. Furlong et al. [31, 36], Walsh et al. [32], and Skinner et al. [34] drew upon methods of ecological niche modelling. However, these habitat suitability models of JEV vectors and reservoir species used static or annual climatic and environmental data and, therefore, were not able to make predictions specific to seasons within a year. Duncan et al. [35] modelled ardeid bird and Culex mosquito habitat suitability on a monthly timescale, with the latter based on a major new Australia-wide collation of existing vector occurrence data. Existing risk assessments that incorporate data on JEV surveillance (e.g., [35]) consist of a limited dataset of published infections [13, 32, 35]. In this work, we collate and describe a dataset of JEV surveillance in humans, domestic pigs, feral pigs, mosquitoes, and sentinel chickens from all Australian states and the Northern Territory, for the period from January 2021 to December 2023. Our dataset included 435 JEV detections and exposures, a five-fold greater number of JEV surveillance records compared to the existing published dataset [35]. Importantly, we also collated a dataset on tests that resulted in JEV-negative records allowing our analysis to account for spatial variation in surveillance effort.

Many approaches to estimating geographic species distributions, including distributions of zoonotic pathogens, are underpinned by ecological niche theory: every species has a set of specific basic requirements to survive in an environment [37]. Within the potential or fundamental niche of a species, all basic habitat requirements are met. The realised niche is a subset of the potential niche, and captures the area where all of a species’ basic environmental needs are met, in the presence of inter- and intraspecies interactions and given spatial and temporal limits on species dispersion. In other words, the realised niche of a pathogen is the area where transmission is occurring.

Here we sought to distinguish between the potential and realised niche of JEV in Australia during the 2021–2023 outbreak. Our analysis constructed these two concepts as follows. To characterise the potential niche of JEV, we used estimates of the distributions of suitable habitat of suspected vertebrate hosts (ardeid birds, feral pigs, and domestic pigs) and mosquito vector species, along with locations of JEV detections, to predict the environmental suitability for JEV transmission: the geographical area that was ecologically accessible to the pathogen at the time of the outbreak (pink, bottom panel of Figure 1). For distributions of suitable habitat for ardeid birds and mosquito vectors, we selected predictions made by Duncan et al. [35] to environmental and climatic conditions in February 2022, the month before the JEV outbreak was declared a Communicable Disease Incident of National Significance, as a representative period when conditions were suitable for JEV transmission. To characterise the realised niche of JEV, we used the locations of JEV surveillance activity (both positive and negative testing records), to estimate the underlying ‘intensity’ of JEV transmission. Because this intensity model does not discriminate between areas with or without environmentally suitable conditions for JEV transmission, we then constrained predictions to geographic areas that are ecologically accessible to the pathogen (i.e., within the potential niche). We considered these areas, where both ecological conditions are met and where there is evidence of recent transmission, to be within the realised niche of the pathogen (red, bottom panel of Figure 1). Outputs from our framework constituted a rapid but nuanced assessment of risk that supported public health responses to the 2021–23 incursion of JEV in Australia.

**Figure 1:**
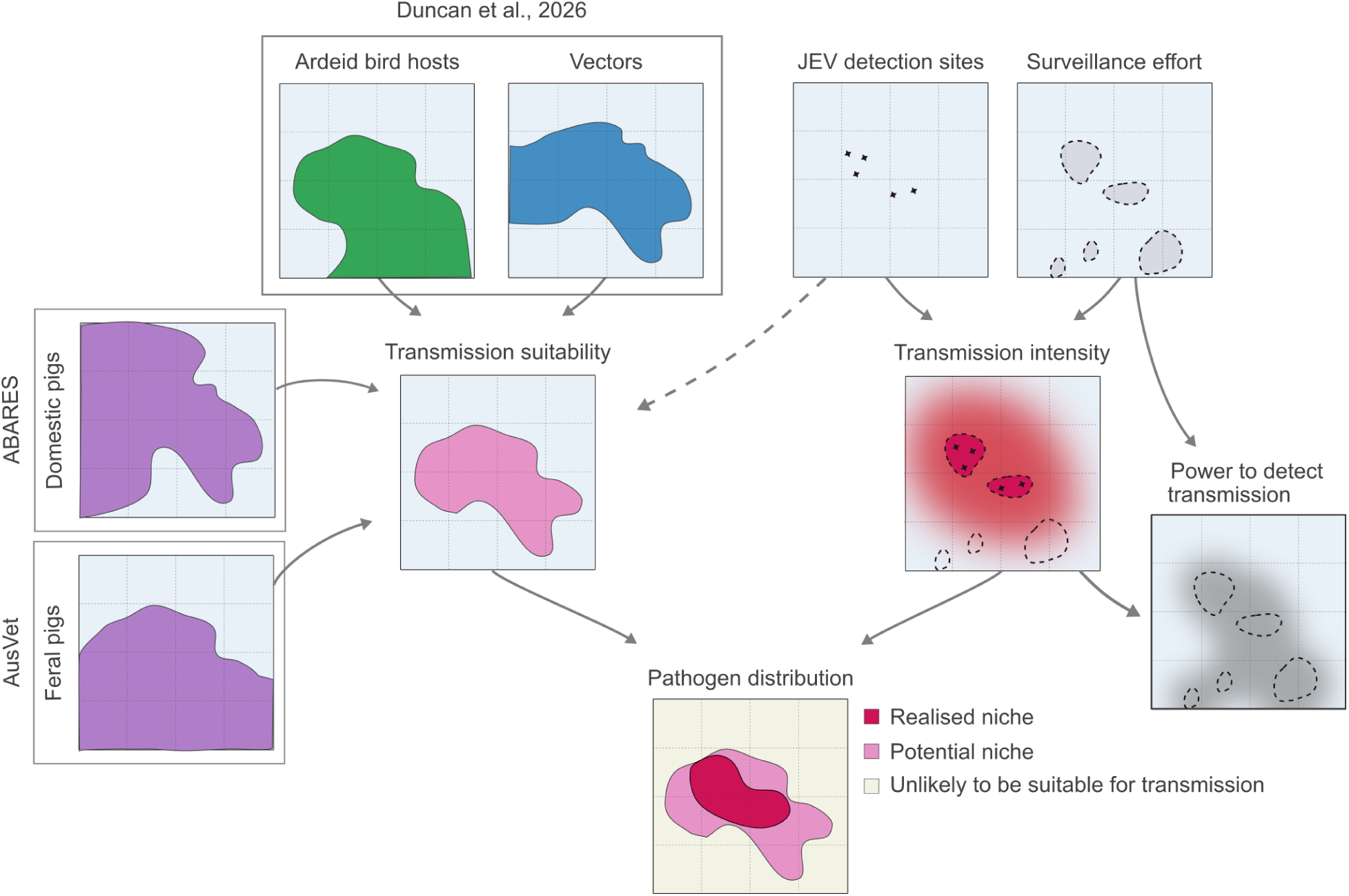
Modelling workflow schematic. Models of ardeid bird hosts (green); mosquito vector species (blue); and maps of commercial pig farms and feral pigs (purple). These host and vector maps are rescaled and combined to estimate JEV transmission suitability (pink). Data of JEV detection and surveillance effort (Table 2) are inputs to a map of JEV transmission intensity during the 2021–23 outbreak (red). Maps of the extent of transmission suitability are used to constrain estimated transmission intensity, to create joint estimates of the extent of the pathogen’s realised and potential niche.

**Table 1:**
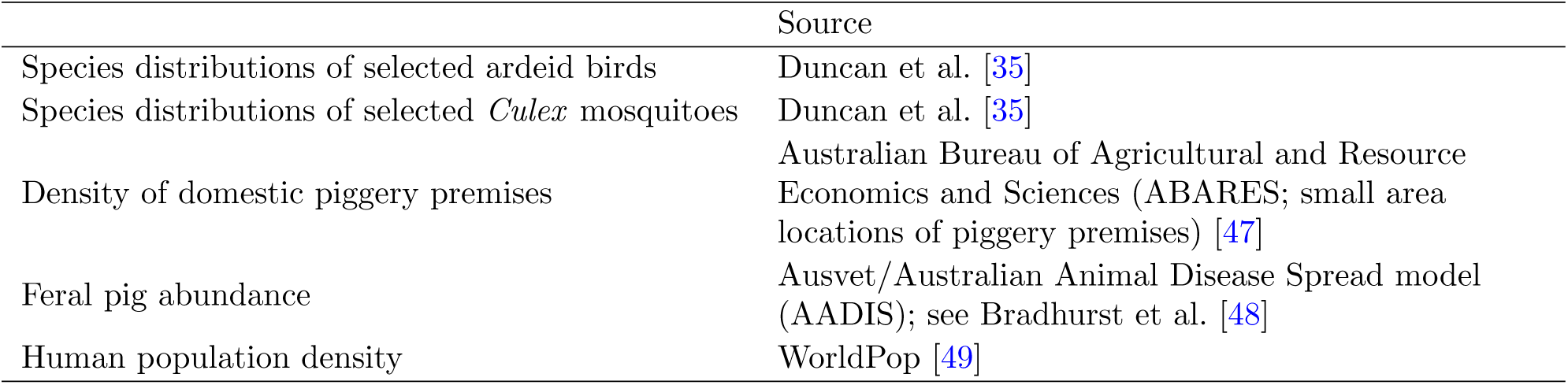
Host and vector density maps and their sources.

**Table 2:** Summary of collated national JEV surveillance/detection data by host and jurisdiction over the period Jan 2021–Nov 2023.

| Host | NSW | Vic | Qld | SA | WA | Tas | NT |
| --- | --- | --- | --- | --- | --- | --- | --- |
| **Humans** - PCR and serological test data |  |  |  |  |  |  |  |
| Cases (n=45)* | 14 | 14 | 5 | 10 | 0 | 0 | 2 |
| Potential exposure sites | 19 | 25 | 16 | 14 | 0 | 0 | 3 |
| **Mosquito trap collections** - PCR test data |  |  |  |  |  |  |  |
| JEV negative | 2,336 | 4,196 | 5,974 | 1,254 | 1,024 | - | 747 |
| JEV PCR positive | 3 | 7 | 4 | 2 | 0 | - | 9<sup>†</sup> |
| **Domestic pigs** - PCR and serological test data |  |  |  |  |  |  |  |
| JEV negative | 70 | 66 | 91 | 32 | 12 | 8 | 3 |
| JEV antibody positive | 27 | 6 | 6 | 4 | 0 | 0 | 0 |
| JEV PCR positive | 29 | 23 | 19 | 10 | 0 | 0 | 0 |
| **Sentinel chicken flocks** - Serological test data |  |  |  |  |  |  |  |
| JEV negative | 187 | - | - | 279 | 273 | - | 25 |
| JEV antibody positive | 1 | - | - | 2 | 6 | - | 0 |
| **Feral pigs** - PCR and serological test data |  |  |  |  |  |  |  |
| JEV negative | 110 | 0 | 475 | 1 | 125 | 0 | 649 |
| JEV antibody positive | 85 | 2 | 8 | 3 | 2 | 0 | 41 |
| JEV PCR positive | 0 | 0 | 6 | 0 | 0 | 0 | 59 |

## Results

### JEV detections and surveillance activities

JEV surveillance activities and clinical investigations conducted by Australian jurisdictions in 2021–23 resulted in 435 detections of JEV or JEV-specific antibodies across humans, domestic pigs, feral pigs, mosquitoes and sentinel chickens. The majority of JEV detections (irrespective of species) occurred between November 2021 and August 2022 in the far north of the Northern Territory and the Murray-Darling basin, including parts of Queensland, New South Wales, Victoria, and South Australia. Two human cases of JEV were recorded in the Northern Territory, with all other human cases (n=43) likely exposed within the Murray-Darling basin, inland of the Great Dividing Range (Figure 2(a)). In mosquitoes, JEV surveillance was generally concentrated around areas of human settlement, including remote communities in tropical latitudes, and areas of domestic pig farming, with detections recorded in northern and central Northern Territory (n=4), the North Burnett region in Queensland (n=4), and within the Murray-Darling basin in New South Wales (n=3), Victoria (n=7), and South Australia (n=2) (Figures 2(b) & 3(b)). Surveillance in domestic pigs covered all areas of the most intensive pig farming (Figures 2(c), 3(c), & S2). Most surveillance effort in feral pigs occurred in northern Australia, particularly near Darwin and coastal areas of the Northern Territory and Cape York, where feral pig destruction and sampling was conducted by the Northern Australia Quarantine Strategy (NAQS) as part of biosecurity operations (Figure 3(d)). Positive JEV samples from feral pigs were reported in the Northern Territory (n=144), Queensland (n=19), Western Australia (n=2), South Australia (n=3), and Victoria (n=2) (Figure 2(d)). At the time of the outbreak, sentinel chicken flocks were situated in South Australia, New South Wales, Western Australia, and the Northern Territory, with JEV exposures reported in flocks in New South Wales (n=1), South Australia (n=2), and northern Western Australia (n=5) (Figure 2(e)).

**Figure 2:**
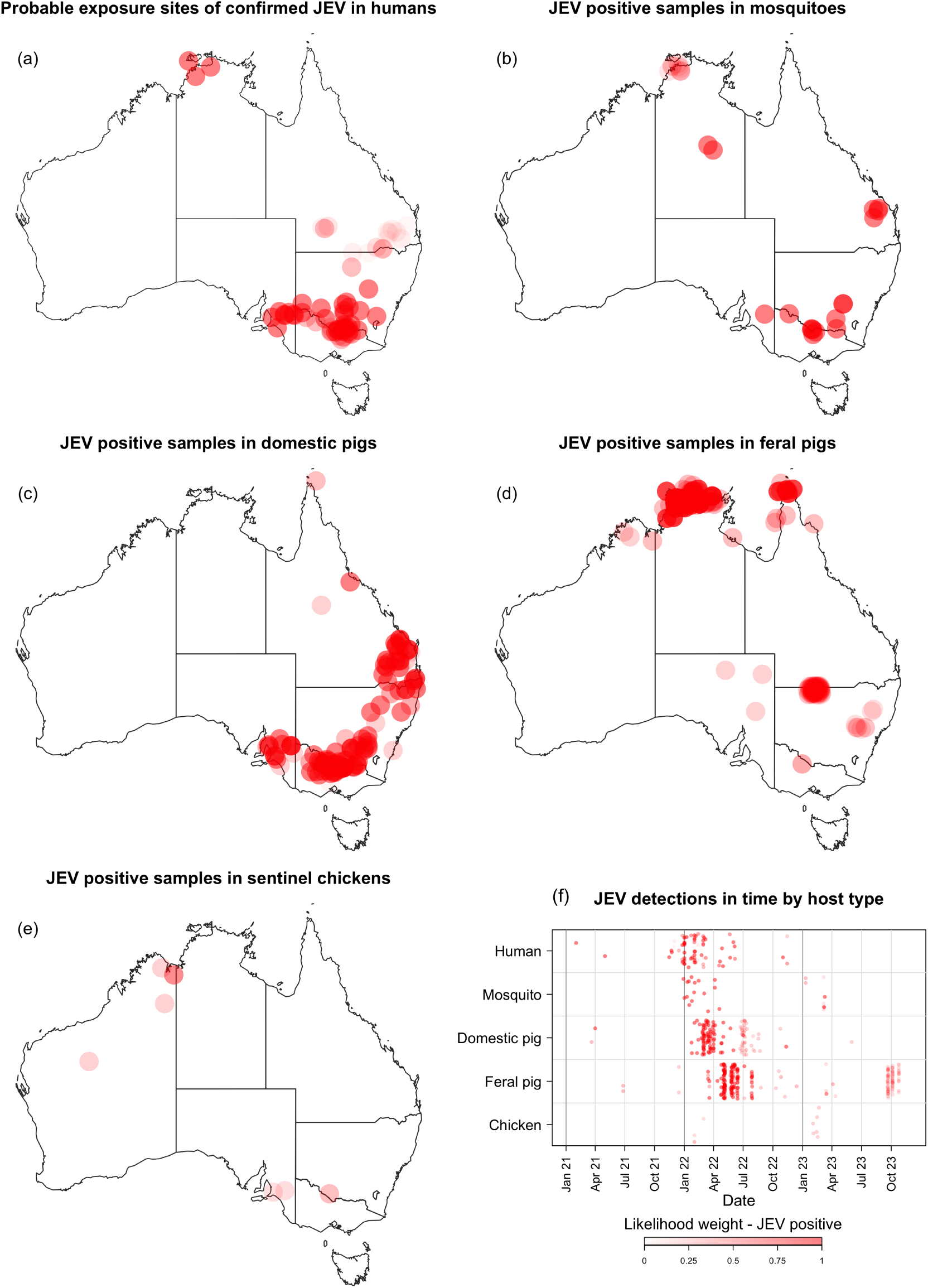
Approximate locations of JEV detections/exposures, with point opacity determined by the diagnostic confidence associated with the surveillance method (Table S1) and the likelihood of JEV exposure at the location, where multiple possible exposure sites were identified for a single exposure. (a) Approximate locations of probable exposure sites for confirmed or probable [38] cases of JEV in humans. (b) Approximate locations of JEV detections in mosquitoes. (c) Approximate locations of JEV detections/exposures in domestic pigs. (d) Approximate locations of JEV detections/exposures in feral pigs. (e) Approximate locations of JEV exposures in sentinel chickens. (f) Distribution of national JEV detections/exposures through time, by host type. Timings and approximate locations of all tests for JEV (with positive or negative outcome) are visualised in Supplementary Figure S10.

**Figure 3:**
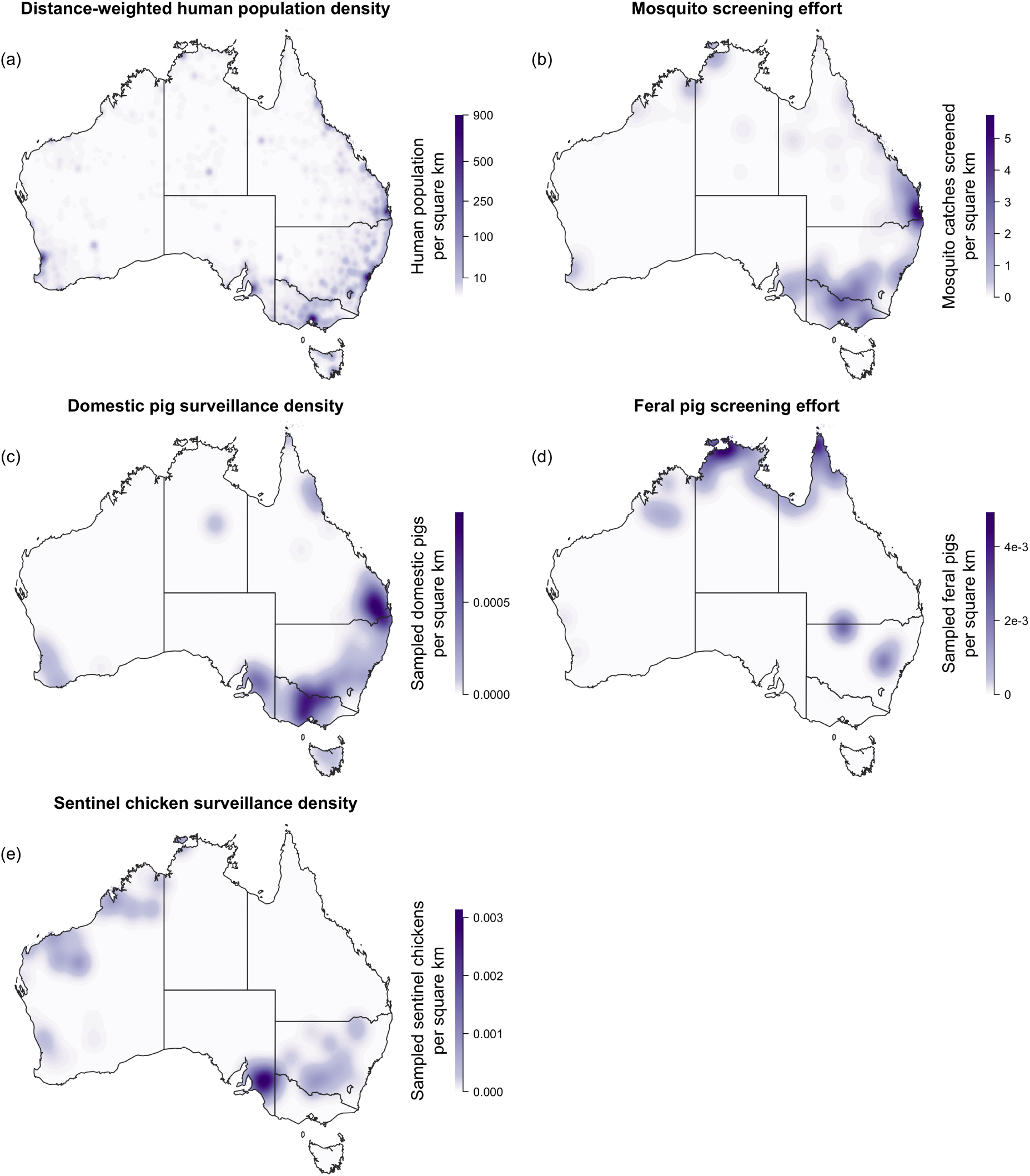
Estimated Australia-wide variation in ability to detect JEV, given 2021–2023 surveillance data in Supplementary Figure S10. (a) Distance-weighted human population density (an indicator of where humans are more likely to be located). (b) Density of mosquito trap catches tested for JEV. (c) Density of domestic pigs or pig premises tested for JEV. (d) Density of feral pig samples tested for JEV. (e) Density of sentinel chickens tested for JEV.

### Potential niche of JEV

When assuming a JEV transmission cycle involving only mosquitoes and ardeid birds (Figure 4(b)), areas of highest predicted suitability for JEV transmission included waterways in the Murray-Darling basin and inland waterways throughout northern areas of Queensland, Western Australia, and the Northern Territory with “summer dominant” rainfall patterns (see Figure 4(a)). Coastal areas of the Northern Territory, north-eastern Western Australia, and western Cape York were also areas of high predicted transmission suitability. Transmission suitability was generally predicted to be relatively low in arid areas, except where they are traversed by significant ephemeral rivers such as those near the South Australia-Queensland border. Tasmania and the Great Dividing Range (from southern Queensland to Victoria) were also areas of low predicted transmission suitability. The inclusion of feral pigs into the transmission cycle (Figure 4(c)) resulted in an increase in predicted transmission suitability across northern Australia, particularly in the Northern Territory and northern Western Australia. With domestic pigs included in the transmission cycle (Figure 4(d)), predicted transmission suitability increased in areas with the highest concentration of pig farming, inland of the Great Dividing Range in eastern Australia (see Figure 4(a); Supplementary Figure S2). Outputs in downstream analyses of our Results use the latter suitability surface, reflecting a transmission cycle including ardeids, feral pigs, and domestic pigs.

**Figure 4:**
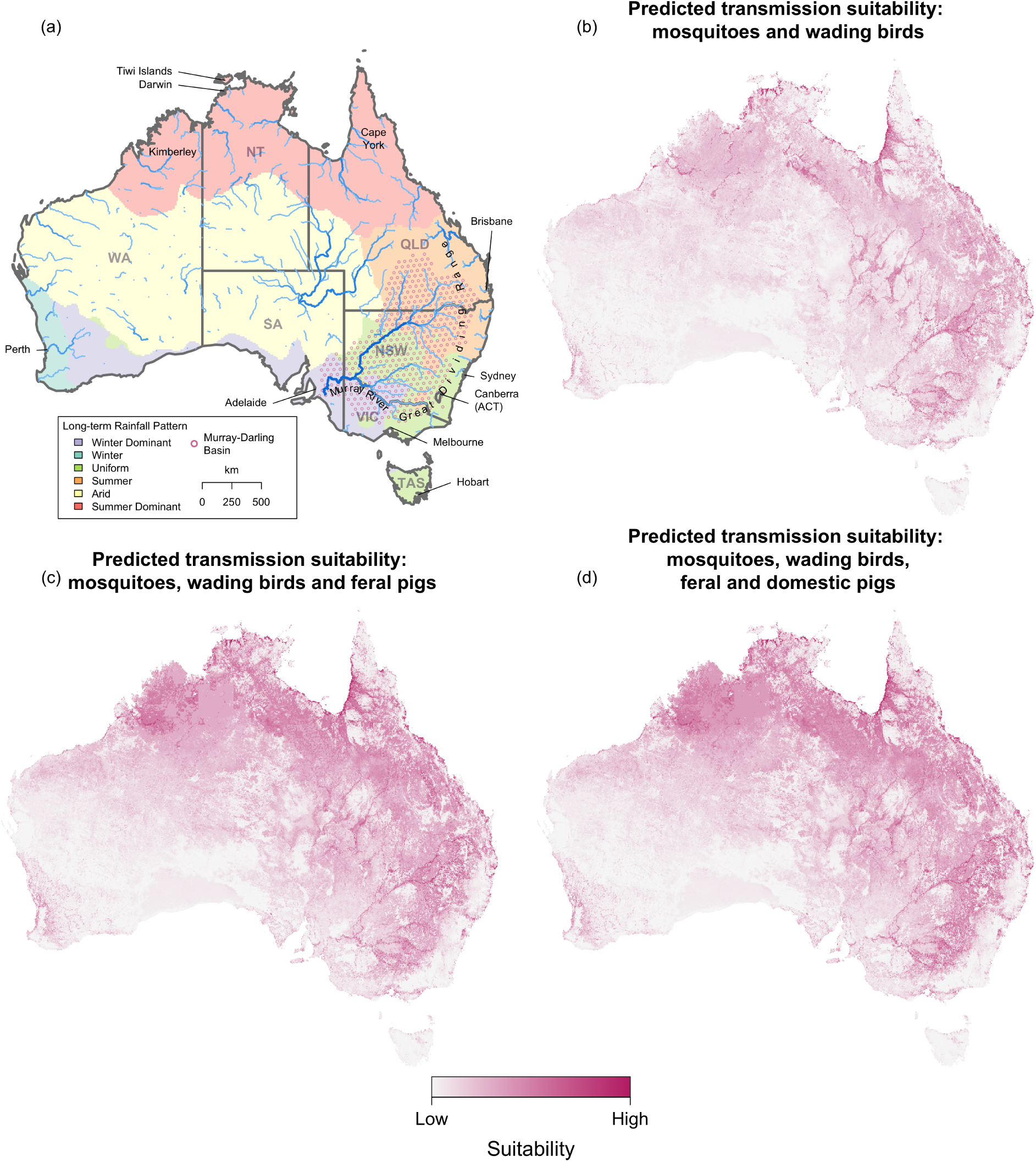
Predicted suitability for JEV transmission in Australia. (a) Map of environmental/administrative landmarks of Australia over map of aggregated seasonal rainfall classifications [39], with approximate scale based on great circle distance at -25^◦^ latitude; predicted suitability for JEV transmission in Australia for February 2022 assuming a transmission cycle including (b) only mosquitoes and ardeid birds (c) mosquitoes, ardeid birds, and feral pigs and (d) mosquitoes, ardeid birds, feral pigs, and domestic pigs. Suitability values are calculated as the combined ‘JEV transmission suitability’ (Section S5), such that habitat suitability or estimated abundance of each component species group is scaled relative to the locations of JEV detections and exposures in 2021–23 (Supplementary Section S2). February 2022 is selected as an example time period, as representative of conditions that preceded the identification of the outbreak: an estimate of transmission suitability for an alternative timeframe is discussed in Supplementary Section S3.

To delineate areas more or less likely to be within the realised and potential niches of JEV, we thresholded our continuous estimates of transmission intensity (see below and Figure 5) and transmission suitability into realised and potential niche extents respectively (Figure 6). When the pathogen’s potential niche (assuming potential transmission in ardeid birds, feral pigs, and domestic pigs) was thresholded to encompass only 50% of all detections and exposures from the outbreak (Figure 6(a)), it included the Kimberley region of north-western Western Australia, and the north of the Northern Territory. Elsewhere, the niche extent tightly tracked waterways and patches of tropical rainforest. Many detections with relative certainty in exposure site (e.g. piggeries where infected animals were fixed in place) were not covered by the niche extent, including in northern Victoria, central New South Wales and South Australia. This threshold therefore resulted in an excessively narrow risk extent: the area within the “true” potential niche was almost certainly more expansive than the area highlighted at the 50% threshold. When 75% of detections are included in the estimated extent of the potential niche (Figure 6(b)), there were substantial areas of transmission suitability across Queensland, northern Northern Territory and north-east Western Australia, as well as the metropolitan areas of Sydney, Melbourne, Brisbane, Darwin, Perth, and Adelaide. The area within the potential niche that includes 95% of detections likely represents an overestimate of transmission suitability, spanning most of Australia, including into large areas of Australia’s arid zone: the area within the “true” potential niche is likely more constricted.

**Figure 5:**
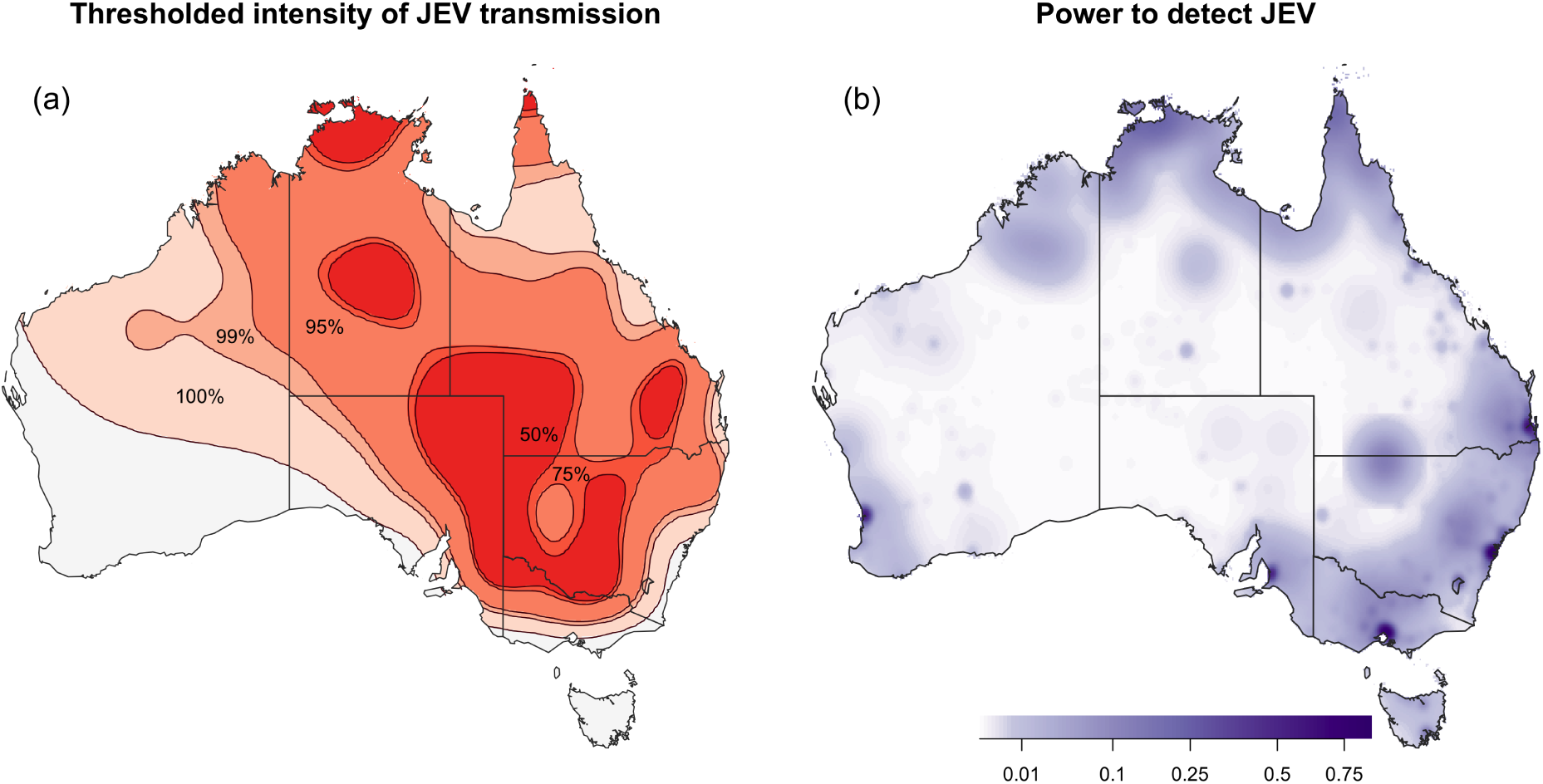
Estimated JEV transmission intensity during the 2021–23 outbreak, estimated from detections in humans, mosquitoes, and feral and domestic pigs, and the overall power to detect JEV via any of these datasets. (a) Possible extents of viral presence according to different thresholds of the continuous intensity surface, i.e., the highest-intensity areas that span 50%, 75%, 95%, 99% and 100% of all JEV detections. (b) The probability of observing one or more JEV detections of any type per (nominal) 11km × 11km map pixel, if it were present at an intensity equivalent to the estimated intensity at the 50% contour in (a).

**Figure 6:**
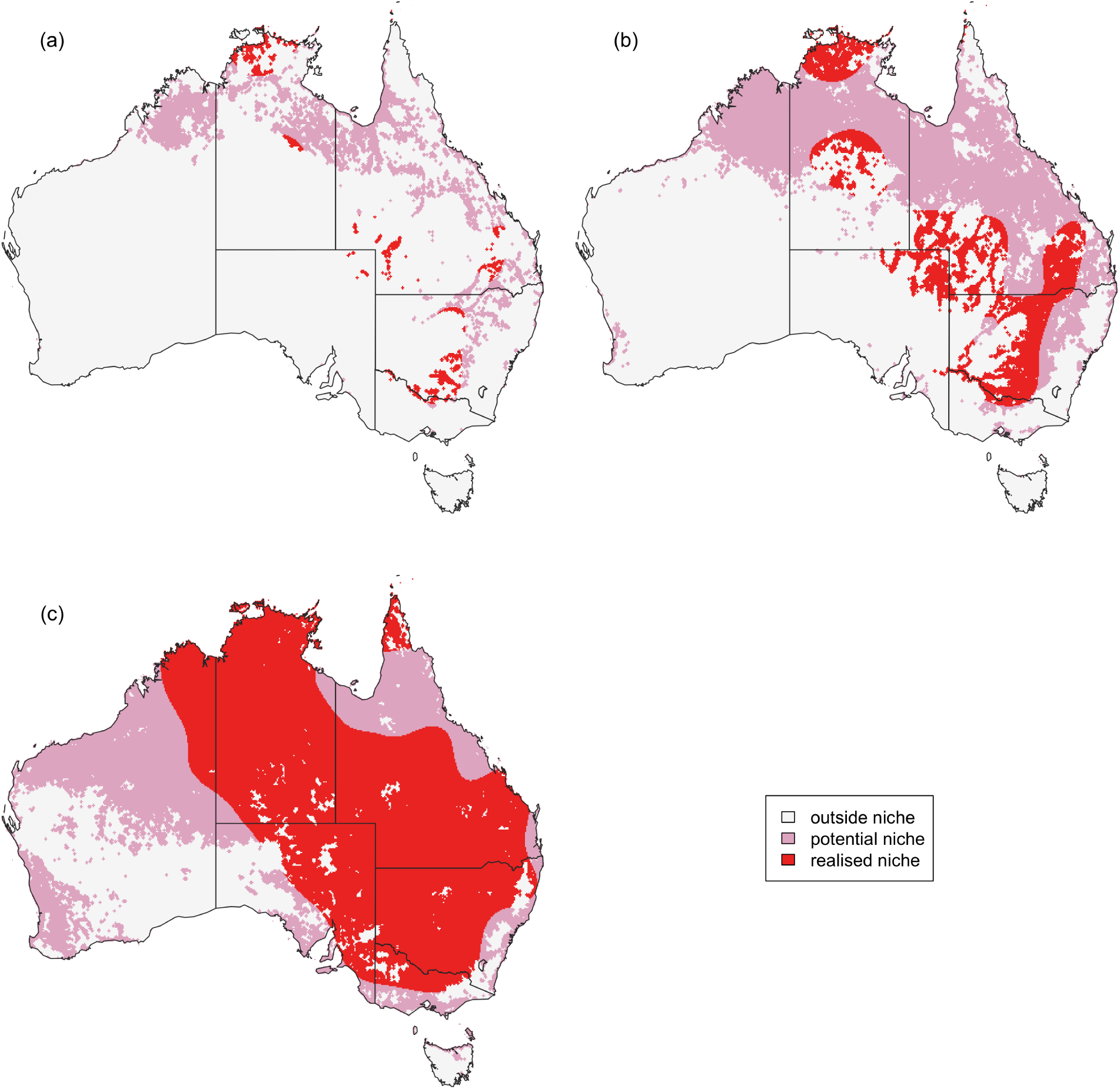
Predicted JEV niche extents. Distributions of predicted JEV transmission intensity (realised niche), and transmission suitability (potential niche) higher than transmission intensity and suitability associated with the locations of (a) 50%, (b) 75% and (c) 95% of all JEV detections. The estimated transmission suitability surface assumes transmission in ardeid birds, feral pigs, and domestic pigs (Figure 4(d)).

### Realised niche of JEV

Using the locations of JEV detections—and accounting for the spatial distribution of surveillance effort, but not the habitat suitability of vector or host species—we estimated JEV transmission intensity during the 2021–2023 outbreak (Figure 5(a)). The following areas were predicted to have the highest intensity of JEV transmission: in the Northern Territory near Darwin and Tennant Creek, in far north Cape York and southern Queensland, in north-eastern South Australia, west of the Great Dividing Range in New South Wales and along the Murray River at the Victoria–New South Wales border.

We constrained the transmission intensity surface by the potential niche surface (i.e., by our estimate of the distribution of habitat suitable for JEV transmission; Figure 4(d)) to generate realised niche extents (Figure 6, in red). Similarly to the potential niche, when the realised niche was thresholded to encompass only 50% of all detections from the outbreak (Figure 6(a)), the estimated extent was almost certainly more restricted than the area within the “true” realised niche. The realised niche extent including 75% of JEV detections spans central and northern regions of the Northern Territory, large areas of south-west Queensland, areas of New South Wales inland of the Great Dividing Range, northern Victoria near the Murray River, and north-east South Australia. The realised niche extent including 95% of JEV detections (Figure 6(c)) spans the majority of eastern Australia, the Northern Territory and northern Western Australia, as well as large regions of desert in central Australia (Figure 4(a)).

### Power to detect JEV

The power to detect JEV in any of the species in our 2021–2023 surveillance dataset (humans, mosquitoes, chickens, and pigs), or the probability of detecting JEV given that JEV transmission was present at high intensity (equivalent to that estimated along the Murray River between New South Wales and Victoria), was estimated to be highest in and around capital cities, where passive surveillance effort (i.e., through symptomatic humans) was high (Figure 5(b)). Estimated power was lowest in remote areas of Australia, away from urban centres and regional surveillance efforts (Supplementary Figure S8). In other words, when holding transmission intensity constant, the estimated probability of detecting JEV transmission was lowest in areas where no surveillance was carried out (as expected). With regards to species groups, per-map pixel power to detect transmission was estimated to be lower in mosquitoes and sentinel chickens (Supplementary Figure S8), which may reflect more complete capture of negative test results (effort) in the surveillance datasets for those groups, relative to humans, domestic pigs and feral pigs.

To visualise spatial variation in surveillance effort by species and to subsequently estimate combined power to detect JEV transmission across species groups, as in Figure 5 and the above results, we used kernel density estimates to represent surveillance effort in non-humans as a smoothed surface where the effort at discrete surveillance locations is distributed across space (Figure 3). When we instead aggregate realised surveillance efforts to Local Government Areas, the probability of detecting JEV, assuming high transmission intensity, was close to one in humans in cities, domestic pigs in areas of intense pig farming in the Murray-Darling Basin, and feral pigs in northern Australia (Supplementary Figure S9).

## Discussion

To characterise the geospatial distribution of JEV infection risk in Australia during the 2021–2023 outbreak, we brought together a highly spatially resolved national dataset of JEV surveillance carried out across 15 state-level and national government departments. Surveillance for JEV was conducted in humans, mosquitoes, domestic pigs, feral pigs, and sentinel chickens across Australia. These data have not previously been collated, harmonised and used in combination to inform high-resolution modelling of geospatial risk. We characterised potential JEV transmission suitability, drawing upon estimates of ardeid bird, domestic pig, feral pig, and Culex mosquito species distribution, where environmental conditions were suitable for presence of the virus, and treated these estimates as expressions of the potential niche of JEV. We then estimated transmission intensity during the 2021–2023 outbreak, which once constrained by our potential niche surface described the realised niche of JEV in Australia at that time. Our maps of the potential and realised niche are presented in terms of ‘risk contours’ to transform the abstract continuous units of the constituent surfaces (e.g., ‘habitat suitability’) to units accessible to decision-makers. The goal of this approach was to enable decision-makers to stratify and prioritise at-risk populations and surveillance locations. The risk contours indicate areas that are relatively more likely to be within the potential and realised niche; they do not provide an absolute quantification of the probability of transmission having occurred or occurring in future.

The estimated extent of JEV transmission during the 2021–2023 outbreak (i.e., the area likely to be within the realised niche) spanned large areas of central and north Northern Territory, south-west Queensland, inland of the Great Dividing Range in New South Wales, northern Victoria near the Murray River, and north-east South Australia. Large areas of eastern and northern Australia also exhibited environmental conditions conducive to transmission of JEV (i.e., likely to be within the potential niche), including several capital cities. However, transmission may not have occurred in these areas during the 2021–2023 outbreak or occur in future outbreaks for several reasons. While the presence of reservoir hosts and vectors is a necessary condition for JEV transmission, the abundance of hosts and vectors in many of these places may well be too low to sustain transmission. Furthermore, geographic barriers separate the capitals of Sydney, Melbourne, Brisbane, Perth, and Adelaide from other areas within the potential niche, including the Great Dividing Range in the east and large areas of desert in the west. If transmission were occurring in major cities, we estimate a relatively high power to detect, given the likely effect of passive surveillance of symptomatic human cases.

In Far North Queensland, we estimated transmission suitability to be relatively low compared to other regions where JEV has been detected, resulting in large omissions from lower-thresholded niche maps. This result was caused by relatively low estimates of habitat suitability in Far North Queensland for either ardeid birds (for a transmission cycle including only ardeid birds as hosts) or Culex mosquitoes (if both ardeid birds and feral pigs are included in the transmission cycle; Supplementary Figure S4). Duncan et al. note that the dataset informing their model of Culex habitat suitability (used in our analysis) lacks sampling of forested habitats, such as those in Far North Queensland, where JEV activity has been detected in historical outbreaks [10].

The Australian landmass is highly variable, encompassing tropical to cool-temperate environments with varying seasonality. The distribution of habitat suitable for JEV transmission will likely contract and expand, particularly in response to cyclic regional drivers of precipitation such as El Niño/ La Niña [32, 40], with great expansion potential during wet La Niña phases which activate expansive ephemeral wetlands in the interior. The estimates of habitat suitability of JEV vector and avian host species produced by Duncan et al. varied temporally [35], with much lower predicted suitability for both species groups in southern Australia in winter compared to summer. A limitation of our analysis is that results are temporally static, informed by Duncan et al.’s habitat suitability predictions for February 2022 and therefore relevant to environmental conditions at that time, coinciding with late summer during a La Niña phase. We describe the potential niche with respect to February 2022 as this period is representative of conditions that preceded the identification of the outbreak. Consequently, our estimates of the potential niche extent are likely maximal, following a hot, wet summer, with the niche likely to contract in southern areas during colder winters and drier summers. Our work could be extended to describe the potential niche during other time periods (and their associated environmental conditions), for example, relating each detection record to a prediction of habitat suitability for the month and year during which the detection was recorded. As an example, we include an alternative, also temporally static, representation of the potential niche of JEV for an average of Duncan et al.’s predictions for the entire 2021–2022 summer in Supplementary Section S3.

We modelled the realised niche of JEV to be constrained by our estimate of the distribution of habitat suitable for JEV transmission. Our approach was therefore able to exclude areas from the realised niche that were proximal to JEV detections if habitat suitability for species involved in the transmission cycle was predicted to be low in these areas. We were also able to distinguish areas where the disease was unlikely to be present (high detection effort, no detection; e.g., in major cities) from areas where the disease may be present but has not been detected (low detection effort, no detection; e.g., western parts of the Murray-Darling basin). Outside of major cities, we estimated the power to detect JEV transmission overall to be low (Figure 5(b)) and the power to detect JEV in humans, mosquitoes, and domestic pigs (Figure 3) was strongly concentrated in areas of high density of domestic pig premises, where surveillance efforts were elevated.

There were several challenges posed by the diversity of hosts, diagnostic methods, and surveillance programs contributing to the national dataset of JEV surveillance. In some cases, uncertainty in exposure site was great: some human cases of JEV were associated with a township or postcode, which generally contain significant environmental variation. We incorporated uncertainty in exposure site by presenting our results (Figures 4, 5 & 6) in terms of cumulative evidence, through thresholding. For example, a location within the 50% contour of transmission intensity is predicted to have higher transmission intensity than at least 50% of all locations where JEV was detected. While some true exposure locations may be mis-located in the dataset, JEV detections in environmental or geographic proximity to each other represent cumulative evidence of JEV transmission suitability or intensity. There was also variation in diagnostic certainty in the surveillance dataset: the interpretation of serology-based tests is complicated by cross-reactivity between JEV and other closely related flaviviruses. We incorporated this uncertainty by assigning “diagnostic weights” to each surveillance record (Supplementary Table S1). These weights were subjective and based on imperfect measures of JEV test sensitivity and specificity—further quantitative evidence is required to support direct comparison between data produced using different diagnostic methods.

Our predictions of the potential distribution of JEV in Australia are approximately concordant with other appraisals of JEV risk produced in response to the outbreak, with some noteworthy distinctions. Furlong et al. [31] and Skinner et al. [34] both estimated highest enzootic risk of JEV transmission in coastal areas: this finding is largely driven by their estimated probability of vector presence [36] (rather than by estimated distributions of ardeids or other hosts), which was itself informed by a limited number of publicly available records of mosquito occurrence. Walsh et al. [32] estimated JEV “outbreak suitability” with respect to domestic pigs (roughly analogous to a transmission cycle including domestic pigs as a limiting component) and predict high suitability west and north of the Great Dividing Range in eastern Australia, in agreement with increased risk in this area in Figure 4(d), relative to Figure 4(c); the former represents the potential niche under the assumption that domestic pigs contribute to transmission, whereas the latter excludes them from the transmission cycle. While domestic pigs are not required for transmission of JEV to occur, transmission in proximity to dense, naive populations of amplifying hosts represents increased risk of transmission to proximal human populations. In previous outbreaks in the Torres Strait, JEV prevention efforts included relocating domestic pigs away from human residences with mixed results [41]. Transmission of JEV has occurred in Australia in the absence of commercial pig farming, for example, in the Northern Territory, and it is possible that feral pigs play an important role in transmission in these areas. In this work, we extend our original method for estimating the potential niche of JEV transmission as described by Duncan et al. [35]—our original implementation used a limited set of published JEV detections from humans and mosquitoes (and did not include feral or domestic pigs), and so the Duncan et al. estimate of transmission suitability is much lower in northern Australia relative to our estimates in Figures 4(c) and 4(d).

While there were significantly fewer detections of JEV in Australia during the summers of 2022–2023 and 2023–2024 than during 2021–2022, suggesting lower levels of transmission, arbovirus transmission continues to be a threat to public health and animal health in Australia [42–44]. High rainfall associated with the La Niña events of 2021–22 and 2022–23 generated conditions conducive to both mosquito and ardeid bird breeding throughout mainland Australia [5, 45]. In 2024–2025, following high rainfall, there were nine human cases of Japanese encephalitis, including five deaths, in eastern states. It is likely that the frequency of extreme La Niña weather events will increase into the future with global climate change [46]. Now that JEV has emerged on the Australian mainland, and likely established itself in wildlife host populations [35], the threat of new outbreaks will likely persist into the future. There are many unknowns in the current understanding of the transmission ecology of JEV in Australia and how it may vary spatially and temporally [5, 31, 32, 34, 35] and the large number of virus detections during the outbreak in relatively temperate areas is of particular concern. Early detection of JEV and other mosquito-borne diseases through surveillance is essential to protect public health and agricultural interests. Limited surveillance capacity and large geographic areas of operation represent key hurdles to surveillance. Ongoing surveillance of JEV, in combination with other arboviruses, will be required to monitor and understand the transmission cycle as it responds to short- and long-term environmental change. The design of surveillance activities, including early warning systems, should be informed by quantitative appraisals of disease risk, such as those presented in this work. It will be important to iteratively update risk predictions and surveillance activities as new data and eco-epidemiological knowledge become available.

## Methods

### Spatial distributions of host and vector species

Estimated spatial distributions of suitable habitat for cohorts of ardeid birds and Culex mosquito species of interest to JEV transmission ecology in Australia have recently been described in Duncan et al. [35]—we used these estimates in our risk assessment (blue and green panels, Figure 1; Table 1). Duncan et al. argued for a conservative cohort model approach for host and vector species on the basis that the wildlife reservoir species and transmission ecology of JEV in Australia are not yet well-defined. The Ardeid model focused on a shortlist of heron and egret species widely distributed in Australia, including Nycticorax caledonicus and some species identified as hosts in Southeast Asia [45]. The Culex model focused on Culex species that are established in Australia and implicated in the transmission of JEV or other flaviviruses, particularly Cx. annulirostris, which is considered the primary vector in Australia [5, 7]. To predict the relative habitat suitability of these groups of bird and vector species, models were informed by a set of environmental covariates (Table S3) related to landscape hydrology (e.g. surface water retention) and land cover (e.g. vegetation), and time-varying (monthly) climate covariates (e.g. rainfall, temperature). Predictions used in the current paper were generated at a spatial resolution of 1 × 1 km^2^ and are specific to environmental conditions experienced in February 2022 [16], although these predictions stand in for predictions made to any relevant time period. In other words, all downstream analysis presented in this paper could be repeated with a set of predictions for an arbitrary month or months—downstream analysis should be repeated with a different set of predictions where it is relevant to a specific policy question or decision.

We further include estimated spatial distributions of domestic pigs, feral pigs, and humans in our geospatial assessment (purple panels, Figure 1; Table 1). These surfaces are described in Supplementary Section S1.

### JEV surveillance data

JEV surveillance data for humans, mosquitoes, domestic pigs, feral pigs, and sentinel chickens were obtained from Australian and State/Territory health and agriculture government departments and harmonised (summarised in Table 2). In Australia, health and agriculture strategy and budgets, including management of communicable diseases, are a competency of state and territory governments, with some nationally coordinated activities across the northern land sea border and arrival ports, i.e., NAQS. Custodians of all surveillance data are listed in Table S4. The majority of the outbreak surveillance dataset represents pathogen surveillance in mosquitoes—most jurisdictions maintain networks of mosquito traps, in some cases developed over the course of decades in response to past outbreaks of mosquito-borne disease (e.g., [50]). Sero-surveillance of sentinel chicken flocks also formed part of active arbovirus surveillance in some jurisdictions during the outbreak, although prior to 2022, these chickens were not routinely tested for antibodies to JEV. Surveillance in humans and, early in the outbreak, in commercial pig farms, was largely in response to clinical symptoms (which in the case of domestic pigs effectively excludes non-farrowing herds) or focused around other detections of JEV. Later, serosurveys of humans and domestic piggeries were undertaken [51, 52], although human serosurvey data were excluded from this analysis, as background exposure to other flaviviruses (or JEV outside of Australia) was difficult to distinguish from local JEV exposure. Surveillance data from feral pigs were abundant: in Australia, feral pigs are invasive and environmentally destructive, and can be sampled and destroyed without permit (in contrast to native ardeid birds). In response to the outbreak, jurisdictions carried out JEV testing on a variety of other feral species and symptomatic domestic animals [53]. However, data received on other host species (e.g., horses, cattle, other livestock or feral species) and pending or inconclusive test results were excluded from our analysis.

The precision in spatial information associated with the supplied surveillance data varied; records were attributed with precise geographical coordinates, truncated coordinates or administrative areas (e.g. townships), often to maintain data privacy requirements. Coarse spatial information was often supplied to maintain data privacy requirements. Where truncated coordinates were provided, the latitude and longitude were truncated at one decimal place (i.e. rounded down), so that a record could be associated with a rectangular area of approximately 11 × 11 km^2^ extending east and south from the coordinate location. Where the area associated with a record was larger than 11 × 11 km^2^, we took the centroid of the area to represent the site of the sample (Table S4).

The surveillance data comprised the results of both serological and nucleic acid-based polymerase chain reaction (PCR) testing (Table 2). Due to the specificity of PCR assays, results from testing are considered to be highly accurate. PCR tests are predominately used to detect virus in mosquitoes [14, 54]. However, JEV RNA is only detectable for a short period of time in humans and other vertebrates due to the relatively short duration of viraemia, which is up to seven days from the day prior to symptoms appearing in humans [14]. For humans and other mammals, PCR tests tend only to be indicated for clinical investigations, with the exception of feral pig surveillance in the Northern Territory, where PCR is used in conjunction with serological assays. All surveillance in sentinel chickens was serology-based.

It is well recognised that the interpretation of JEV serological test results is complicated by cross-reactivity with antibodies of closely related flaviviruses (e.g., Murray Valley encephalitis virus (MVEV) and the Kunjin strain of West Nile virus (WNV_KUNV_)) [14, 54, 55]. The preferred reference standard for serological testing is the plaque-reduction neutralisation test (PRNT) [56, 57]. Not all samples in the dataset had been subject to tests of equivalent specificity and sensitivity, or the ideal PRNT confirmation. As a result, diagnostic confidence was not constant in our collated dataset of JEV surveillance and we instead constructed varying levels of diagnostic confidence and incorporated them into our model (Supplementary Section S4).

### Potential niche of JEV

To represent the distribution of the potential niche of JEV, we considered areas where there is suitable habitat for Culex mosquitoes, ardeid birds, feral pigs, and domestic pigs. We extended the approach to combining modelling outputs for Culex and ardeid species presented in Duncan et al. [35], who rescaled and combined ardeid host and vector surfaces to find intersecting areas where suitable habitat is present for both hosts and vectors. In our analysis, we also included estimates of feral and domestic pig distributions since they are widely regarded as important amplifying hosts of JEV, albeit not strictly limiting components of the JEV transmission cycle [27]. The maps of relative habitat suitability of Culex mosquitoes and ardeid birds first described in Duncan et al. represent different units: to make the habitat suitability metrics comparable and interpretable in terms of suitability for JEV transmission, we extracted each model’s output at the set of locations where JEV was detected (Table 2) and rescaled the vectors of map values using an empirical cumulative distribution function [35, 58] (Supplementary Section S2). The values in the rescaled maps then corresponded to the cumulative proportion of all JEV detections falling in areas with a given suitability value or lower. For example, 50% of JEV detection locations had a scaled value of 0.50 or lower, 75% of JEV locations had scaled values of 0.75 or lower (and 25% had values above 0.75), and so on. The set of JEV detection locations used in this work differed from those used by Duncan et al.; they drew on an existing set of published detections, as used by Yakob et al. [13, 59] that contained only 19% of the detections in our dataset, was limited to February–April 2022, and did not include evidence of virus exposure in the Northern Territory or Western Australia. This set of detections was published relatively early in the outbreak, before exposures in these jurisdictions had been identified/publicised.

To create a map of overall suitability for a transmission cycle of vectors and bird hosts only, we combined scaled surfaces for mosquitoes and ardeid birds by taking the minimum of the two scaled surfaces, for each pixel. The resulting JEV transmission suitability map therefore shows an intersecting area where both components of this assumed transmission cycle have suitability above a given threshold, i.e. the potential niche of JEV (pink panel, Figure 1).

To visualise overall JEV transmission suitability for transmission cycles including feral and/or domestic pig hosts as well as ardeid birds, we also generated maps of JEV transmission suitability including similarly scaled maps of feral and domestic pig distributions. For transmission cycles involving multiple hosts, we took the maximum of the scaled suitability surfaces across hosts. Though pigs are usually identified as amplifying hosts of JEV [11, 54], we avoided specifying an arbitrary amplifying factor. The transmission suitability surfaces under three possible conceptualisations of the JEV transmission cycle were calculated as follows:

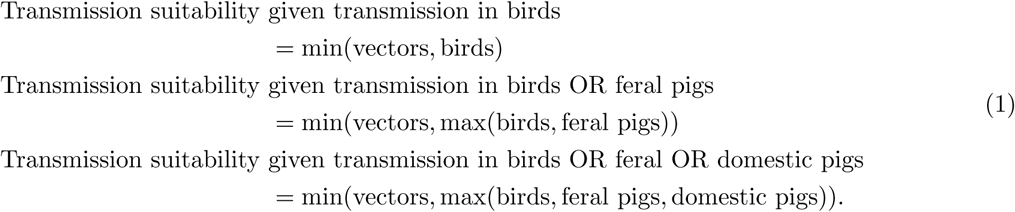

### Realised niche of JEV and overall JEV surveillance effort

In order to estimate the realised distribution of JEV in Australia during the 2021–2023 outbreak (or the area where JEV was likely to have been circulating) we first predicted the underlying ‘intensity’ of JEV transmission (red panel, Figure 1). Our model of transmission intensity explicitly distinguished between the spatial variation in the underlying intensity of JEV risk and in the surveillance ‘effort’, or the probability of detecting JEV in a particular location given it was present, by drawing on our set of spatially and temporally resolved data of JEV surveillance between January 2021–November 2023 (both JEV-positive and -negative records; Table 2).

In this model, we assumed that the intensity of JEV transmission was proportional to: the rate of human detections relative to the estimated surveillance effort; the fraction of mosquito trap collections containing JEV-positive mosquitoes; the fraction of tested pig premises with JEV-positive animals; the fraction of tested feral pigs that were JEV-positive; and the fraction of tested sentinel chickens that were JEV-positive. However, we assumed that constants of proportionality were different for each of these types of JEV detection, and a priori unknown, allowing for differences in underlying infection prevalence and incidence, and infection ascertainment between each species and dataset. Whilst the available data were insufficient to precisely estimate epidemiological parameters, incidence and prevalence data for each species can be assumed to be approximately proportional to the force of infection into that species in a given location. The intensity surface we modelled was therefore a general proxy for the amount of JEV transmission expected to be occurring.

For JEV detections in tested mosquito collections, domestic pigs, feral pigs and sentinel chickens, surveillance effort was measured by the number of samples tested per pixel. In the absence of negative JEV test result data in humans, we modelled the JEV surveillance effort in humans as proportional to the distance-weighted human population density in each pixel. At each pixel, the distance-weighted population was the weighted average of the resident population in all pixels in the surrounding area, with weights decreasing with increasing distance according to a Gaussian function with standard deviation σ = 0.2 decimal degrees (approximately 22 km). This distance-weighted population represented a proxy for the average number of people within a given pixel, who could have been exposed there. Human disease detection data were associated with suspected exposure locations (as determined by public health teams), rather than residences, although our approach assumes that people are more likely to have been exposed close to their residence than far from it.

We accounted for the varying reliability of different diagnostic testing procedures by assigning a subjective ‘weight’ to each type of diagnostic result (both positive and negative) given the relative strength of the evidence provided by a sample of each type. Our proposed schema of weights for all combinations of non-human host types and diagnostic results is shown in Table S1. In brief, the results of PCR tests were assigned a weight of one, with weights equal to or less than one for serological results. Further details are included in Supplementary Section S4.

The observation model for the number of detections, y_i,g,d_ , at each location, i, in each detection group, g (either mosquitoes, humans, domestic pigs, feral pigs, or chickens), of diagnostic type, d (either PCR, serology, or unknown: see Supplementary Section S4), is Poisson, with the expected rate given by the product of the dataset-specific observation intensity surface z_i,g,d_ and the surveillance effort for that group φ_i,g_ . This Poisson formulation is an approximation to the usual binomial distribution for prevalence data. Using the Poisson approximation enables us to combine multiple datasets in a single model, and given the relatively low prevalence of infection in all of these datasets, it is a near-perfect approximation. The logarithm of the dataset-specific observation intensity surfaces were modelled with a penalised isotropic thin-plate spline on geographic space (giving the logarithm of the combined intensity surface), with intercept terms for the detection group, β_g_ , and diagnostic type, γ_d_ , to model the differing constants of proportionality. The positive constants of proportionality for the detection group and diagnostic type were drawn from parameter vectors β and γ. Parameter vectors β and γ were modelled as fixed effects. While there may have been an advantage to inference in modelling β as a random effect with common mean and variance, in practice we found that species-level differences in surveillance efforts and detection rates required the model to have the flexibility of a fixed effect. Hence, the model can be written as:

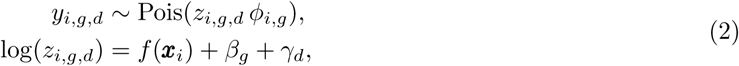

where the model covariates, **x**_i_ , are longitude and latitude.

We fitted the model at a nominal 11 km spatial resolution such that the number of JEV detections (zero in pixels with no JEV detections), and the surveillance effort, were summed over the area of each approximately 11 × 11 km^2^ pixel. We fitted the model as a generalised additive model with restricted maximum likelihood using the R package “mgcv” [60]. We used likelihood weights to represent uncertainty in JEV exposure location and/or diagnostic method. All likelihood weights were less than or equal to one, and assigned to the product of “diagnostic weight” (Supplementary Table S1) and “exposure weight”: where multiple possible exposure locations were provided for a single infection, the total exposure for the infection was allocated across locations, such that the exposure weight for all locations associated with a single infection summed to one. For some human JEV exposures, public health unit staff provided scores indicating the relative likelihood of exposure at each location based on information from case interviews.

Finally, to reach an estimate of the realised distribution of JEV during the Australian outbreak during 2021–2023, we intersected our surface of JEV transmission intensity with our surface of JEV transmission suitability, given transmission in ardeid birds and feral and domestic pigs. Because our transmission intensity surface is based on a space-only model (i.e., associations between transmission intensity and environmental factors are not modelled), this step allows us to constrain transmission intensity to areas where minimal environmental conditions for transmission are met. In other words, a degree of predicted environmental suitability for transmission was assumed to be a limiting constraint on realised transmission intensity. We considered this constrained surface of transmission intensity to represent the realised niche of JEV transmission (bottom panel, Figure 1). By including this step, we were able to exclude areas from our realised niche estimate that are geographically close to but environmentally distant from virus detections.

### Estimating the power to detect JEV transmission

To contextualise our surface of JEV transmission intensity during 2021–2023, we produced maps of statistical power to detect JEV. We considered statistical power to detect JEV in humans, mosquitoes, domestic pigs, feral pigs and sentinel chickens independently and for all five groups combined. These maps represent the probability that a JEV detection of any type would occur in a given pixel, given available data of surveillance activities between 2021–2023, if JEV transmission were present at relatively high intensity. Statistical power for each detection type was calculated as the probability of observing one or more JEV detections under a Poisson sampling assumption (Eq (2)), incorporating the surveillance activities for that detection type, and assuming that the underlying intensity was at the level corresponding to the 50% threshold extent (i.e., assuming the underlying transmission intensity was relatively high: 50% of JEV detections were associated with an estimated transmission intensity below the threshold). To compute the overall power across all detection types, we combined the component probabilities, assuming conditional independence. The calculation of power-to-detect outputs is described in detail in Supplementary Section S5.

To aid in visualisation of the effort from these surveillance activities when viewed at a national scale, we calculated a map of smoothed surveillance effort using a two-dimensional Gaussian kernel density estimate of surveillance locations.

### Defining potential and realised niche extents

The estimated transmission suitability and constrained transmission intensity maps are on a continuous scale, indicating areas that are more likely to be within the potential and realised niches of JEV transmission, respectively. In order to prioritise geographic regions and their populations for surveillance and interventions, we arbitrarily classified these outputs into discrete levels of risk. We formed “contours” of transmission suitability or intensity where all locations with suitability or intensity at or above a threshold value are included in the corresponding contour and risk area. In order to define interpretable contours, as before, we computed suitability and intensity thresholds relative to the proportion of all JEV detection locations included within the given contour. For example, to define a 50% threshold region of potential niche suitability, we found the median (i.e. 50th percentile) suitability of all locations where a JEV detection was assigned, then considered all pixels with this suitability or higher to fall within the 50% threshold region. We first applied these thresholds separately to the transmission intensity surface and to the transmission suitability surface, then intersected the thresholded transmission intensity region with the corresponding transmission suitability region to estimate the realised niche.

The construction of our outputs around thresholds allows us to visualise the cumulative evidence for an area falling within the potential or realised niches. All threshold outputs should therefore be interpreted together to gain the best understanding of where this cumulative evidence is greatest and sparsest. The thresholding approach also allows us to account for measurement error: we assume that all true transmission locations should fall within both the potential and realised pathogen distributions, but that some fraction of detections might be recorded at a location that is not the site of transmission and therefore may not be within the true extent of transmission. For example, for some human detections, the residence location may be given instead of the place of actual exposure; when we assign a detection associated with a larger polygon to the coordinate of a single map pixel, the environment at this pixel may not be within the true potential niche.

## Code availability

All code is available at https://github.com/lu-harr/jev_aus_risk.

## Data Availability

All data produced in the present study are available upon reasonable request to the authors.

## Acknowledgements

The authors thank the many local governments and personnel who support JEV-based surveillance activities in Australia. We would also like to acknowledge the following individuals, teams and departments who provided data and/or assistance in its interpretation:

1. New South Wales Health and Health Pathology New South Wales (through Department of Medical Entomology)
2. New South Wales Department of Primary Industries and Regional Development Virology Laboratory and the Animal Biosecurity Unit at the Elizabeth Macarthur Agriculture Institute (EMAI)
3. Andrew Bengsen from the Vertebrate Pest Research Unit, New South Wales Department of Primary Industries and Regional Development
4. Northern Territory Department of Agriculture and Fisheries,
5. Northern Territory Health Medical Entomology team,
6. Northern Territory Centre for Disease Control, including Dr Vicki Krause,
7. Queensland Public Health Units,
8. Population Health Division of Queensland Department of Health,
9. Public and Environmental Health Reference Laboratories (PEHRL), Pathology Queensland,
10. Queensland Department of Primary Industries, including Dr Joanne Mollinger, Dr Selina M. Ossedryver, and the Biosecurity Sciences Laboratory,
11. Communicable Diseases Control Branch, South Australia Health
12. Department of Primary Industries and Regions South Australia
13. Tasmanian Department of Health,
14. Julia Sarandopoulos, Dr Leanna Dries, Dr Sarah Hall, and Leah Starick from Agriculture Victoria, Department of Energy, Environment and Climate Action
15. Community and Public Health Division, Victorian Department of Health, in particular Dr Rebecca Feldman and A/Prof. Jim Black
16. Western Australia Health Medical Entomology Team,
17. Western Australia Department of Primary Industries and Regional Development, Animal Biosecurity & Welfare,
18. Commonwealth Scientific and Industrial Research Organisation (CSIRO) Australian Centre for Disease Preparedness (ACDP)
19. Northern Australia Quarantine Strategy (NAQS)
20. Dr Rachel Iglesias from the Office of the Chief Veterinary Officer, Department of Agriculture, Fisheries and Forestry

Thanks also to Associate Professor Simon Firestone, Professor James McCaw, Professor Bart Currie, and members of the Australian National Arbovirus and Malaria Advisory Committee (NAMAC) and the Communicable Diseases Network of Australia (CDNA) for expert advice and feedback during development of the modelling framework. Thanks to Dr Domenic Germano for assistance in obtaining and interpreting incoming data.

## Author contributions

Conceptualization: FMS and NG. Data curation: LEH, DHD, XT, PJN, CCJ, AP, CB, KLM, MH, DTW, and SLD. Formal analysis: LEH, DHD, FMS, and NG. Funding acquisition: FMS. Investigation: LEH, DHD, FMS, NG, and DTW. Methodology: LEH, FMS, NG, and CCJ. Project administration: DHD and FMS. Resources: FMS, NG, PJN, DTW, and SLD. Software: LEH, DHD, FMS, and NG. Supervision: FMS, NG, DJP, and JAF. Validation: LEH, DHD, and PJN. Visualisation: LEH, FMS, and NG. Writing - original draft: LEH. All authors contributed to Writing - review & editing.

## Data availability

We were unable to obtain permission from the institutions to publicly archive the data collated for this study due to concerns about the sensitivity of surveillance locations on commercial and private property, however, the component datasets can be requested from each institution detailed in the Acknowledgements section via a data sharing agreement.

