## Supplementary material for "Geospatial assessment of Japanese encephalitis virus infection risk in Australia"

#### Contents

**S1 National maps of non-ardeid hosts** **1**

**S2 Notes on vector and host map scaling** **1**

**S3 Transmission suitability predictions using alternative habitat suitability timeframe** **6**

**S4 Notes on serological testing and diagnostic uncertainty** **8**

**S5 Detailed description of power to detect JEV transmission** **13**

**S6 Further supplementary figures and tables** **16**

### S1 National maps of non-ardeid hosts

Together with estimates of *Culex* vector and ardeid host distribution, we include estimates of the spatial distributions of commercial pig farms, feral pigs, and human population density into our modelling framework (Fig 1). A national dataset containing the verified locations and numbers of domestic pigs and commercial pig farms was not publicly available at the time of analysis. We therefore generated a surface of *pig farm density* from a national dataset containing small-area locations of pig farms [1]. This dataset contained polygons representing 1,257 sites, documented between 2008 and 2019. There was considerable duplication and overlap in this dataset: by merging polygons that share at least one edge, we reduced the number of instances in the dataset from 1,257 to 840. We then created a smoothed map from the centroids of pig farm polygons, via a two-dimensional Gaussian kernel density estimate with standard deviation, $\sigma$, of 0.4 decimal degrees [2] (Supplementary Figure S2(a)).

A national map of feral pig abundance (Supplementary Figure S2(b)) was previously published by AusVet to inform the Australian Animal Disease Spread (AADIS) model [3]. The map was based on available occurrence data up to 2008 [4] and an estimate of national abundance generated in 1990 [5]. There are known limitations to this map and an improved model of feral pig abundance was published by the Commonwealth Department of Agriculture, Fisheries and Forestry in 2024 [6]. However, we include the AusVet model in this work as it was the best available quantitative estimate of the spatial distribution of feral pigs in Australia at the time of our analysis. A surface of human population density was accessed from WorldPop [7]. All contributing host and vector density maps are summarised in Table 1.

### S2 Notes on vector and host map scaling

Maps of estimated *Culex* mosquito habitat suitability and ardeid bird habitat suitability relevant to February 2022, as described in Duncan et al. [8] are attached in Supplementary Figures S1(a) and S1(b), respectively. These surfaces subsequently scaled by empirical cumulative distribution functions of the locations of JEV detections are attached in Supplementary Figures S1(c) and S1(d). Estimates of the spatial distributions of domestic pig premises and feral pigs are attached in Supplementary Figures S2(a) and S2(b), respectively.

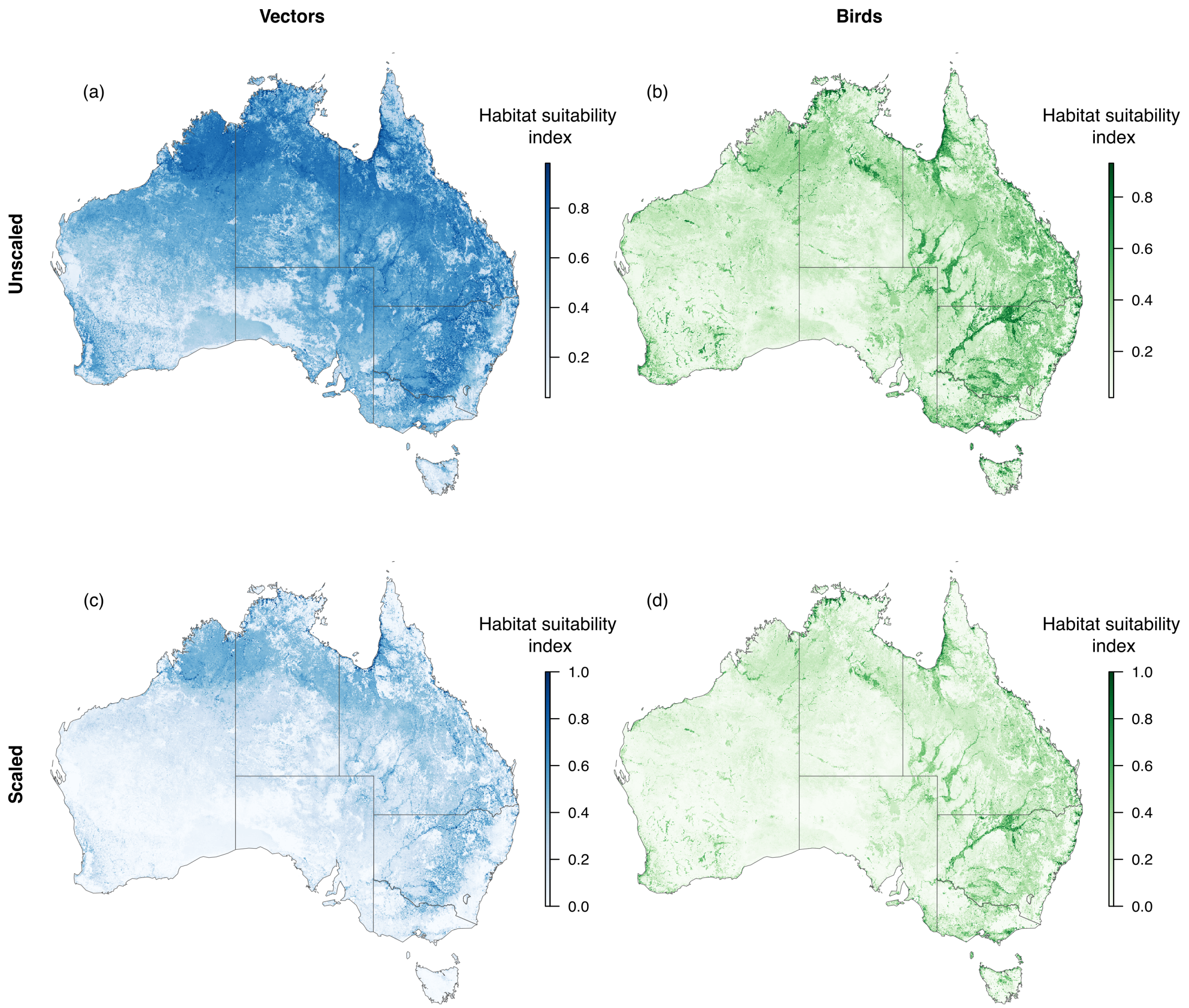

**Figure S1: Estimated habitat suitability of *Culex* mosquitoes and ardeid birds in Australia in February 2022**. (a) and (b) show estimated habitat suitability of *Culex* mosquitoes and ardeid birds in Australia in February 2022 as described in Duncan et al. [8]. The maps in (c) and (d) are the habitat suitability estimates in (a) and (b) following scaling by empirical cumulative distribution functions of the locations of JEV detections.

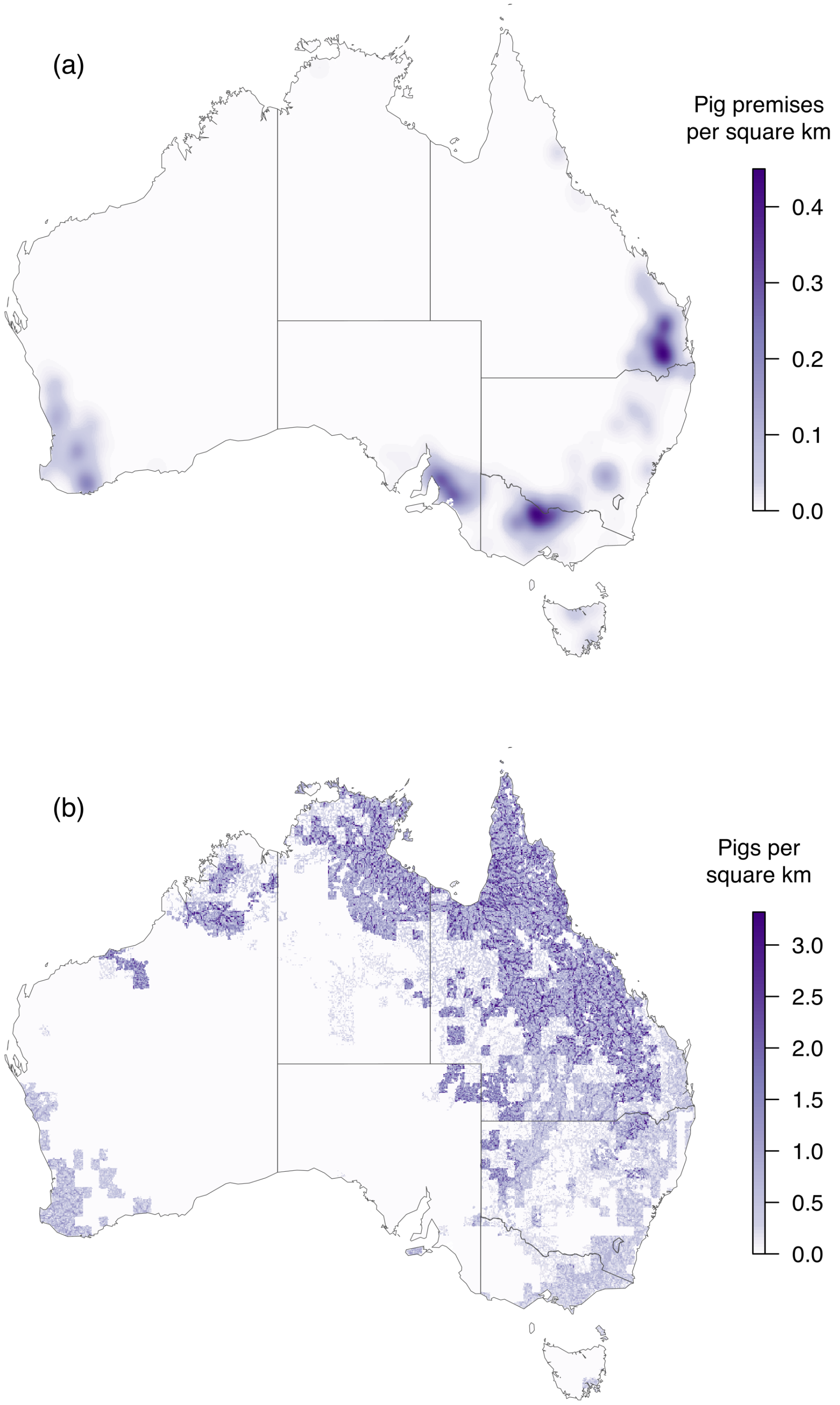

**Figure S2: Estimated distributions of domestic pig premises and feral pigs in Australia.** (a) Density of domestic pig premises estimated from the set of small-area locations of domestic piggeries published by ABARES [1]. (b) Estimated abundance of feral pigs [3], given national estimates of abundance [5] and limited observations [4].

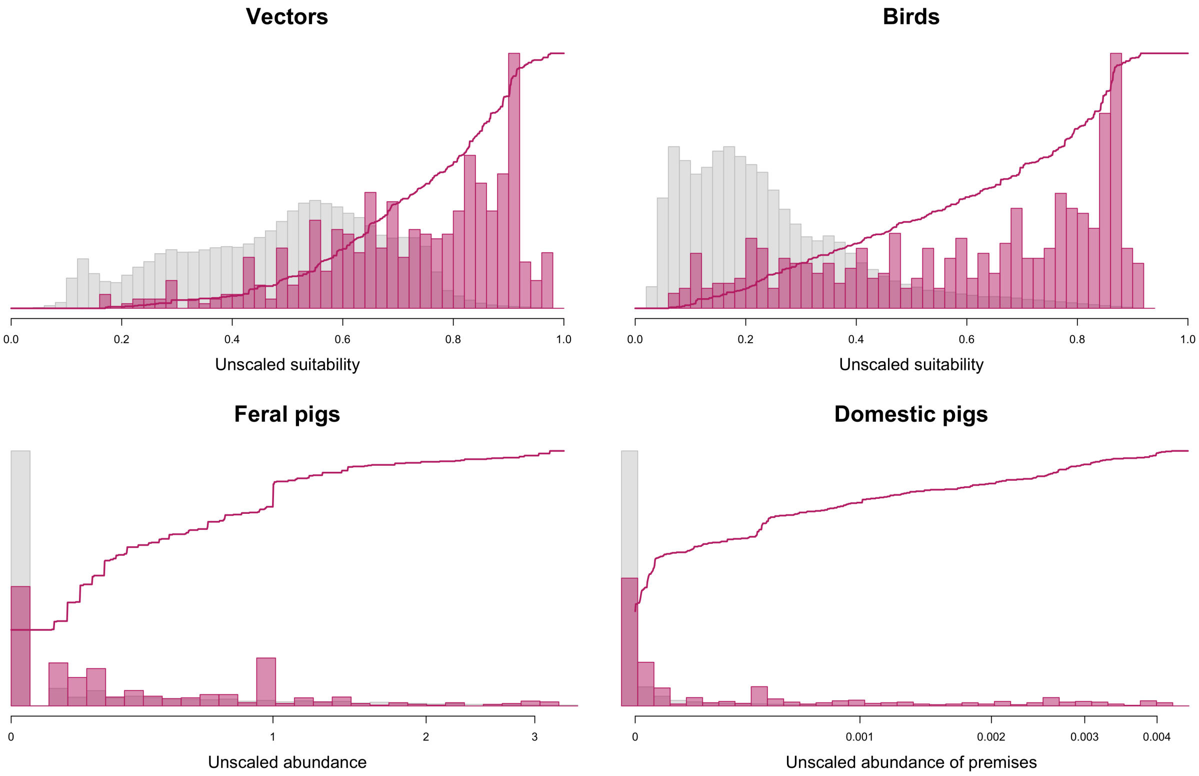

**Figure S3: Empirical cumulative distribution functions of species distribution maps at locations of JEV detections.** Histograms of all values in distribution maps (grey) and values in distribution maps associated with JEV detections (pink), with empirical cumulative distribution function of JEV detections superimposed for (a) *Culex* mosquito vectors, (b) ardeid birds, (c) domestic pigs, and (d) feral pigs.

Empirical cumulative distribution functions of the set of vector and host map values (e.g., as applied in Supplementary Figure S1) are included in Supplementary Figure S3. Broadly speaking, the mapped habitat suitability/density/abundance of each species group is higher at locations of JEV detections (pink) compared to the overall map (grey). There are detections of JEV at locations where estimated abundance of feral pigs and estimated density of domestic piggeries is zero (see Figure S10): neither of these groups are limiting components of the transmission cycle.

The constituent surfaces contributing to potential niche estimates in Figure 4 are visualised in Supplementary Figure S4. Under a transmission cycle involving only *Culex* vectors and ardeid hosts (Figure 4(b)), the majority of the transmission suitability surface is 'controlled' by the ardeid surface, compared to the mosquito surface (Supplementary Figure S4(a)). In other words, the scaled habitat suitability for ardeids is less than the scaled habitat suitability for mosquitoes in the majority of locations, particularly northern, western and central Australia. When feral pigs are also included into the transmission cycle, the joint relative suitability for hosts (i.e., *birds OR feral pigs*) is generally greater than the relative suitability for mosquitoes: because of the minimum operation in Eq (1), the majority of the values in Figure 4(c) are controlled by values in the mosquito suitability surface, with the exception of parts of the Kimberley region in northern Western Australia, and southern parts of the Victoria-Daly Shire and eastern Central Desert region in the Northern Territory (Supplementary Figure S4(b)). The inclusion of domestic pigs into the transmission cycle (Figure 4(d)) further increases the joint relative suitability for hosts (*birds OR feral pigs OR domestic pigs*): there are more blue pixels in Supplementary Figure S4(c) relative to Supplementary Figure S4(d), particularly in areas where relative suitability for domestic pigs is high, near the Great Dividing Range and in south-west WA. These areas are highlighted in the host-only map (*birds OR feral pigs OR domestic pigs*), visualised without a constraint on mosquito habitat suitability in Supplementary Figure S4(d).

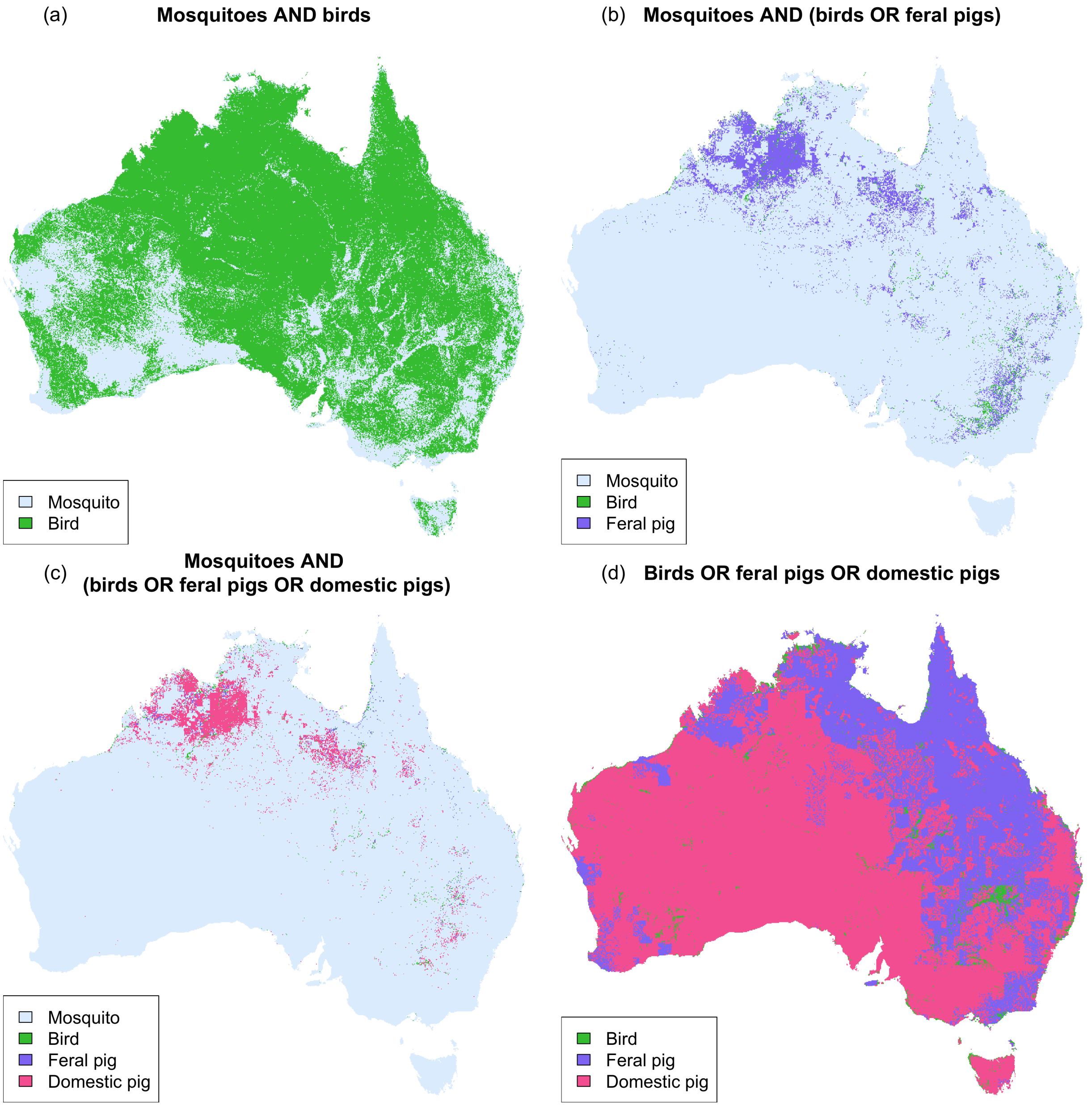

**Figure S4: Contribution of distribution maps of *Culex* mosquitoes, ardeid birds, feral, and domestic pigs to maps of potential niche.** In (a), colour at each pixel indicates whether scaled suitability of ardeid birds is less than scaled suitability of *Culex* mosquitoes. In (b) and (c), colour at each pixel either indicates that scaled suitability for *Culex* mosquitoes is less than scaled suitability for non-human hosts, or the host with greatest scaled suitability. Panels (a), (b), and (c) correspond to transmission cycles visualised in Figures 4(b), 4(c), and 4(d), respectively. In (d), the contributions of hosts to the transmission cycle in (c) are visualised in the absence of vectors as a limiting constraint (this is not a viable transmission cycle, but is included to highlight relative contributions of hosts in (c)).

### S3 Transmission suitability predictions using alternative habitat suitability timeframe

Within the main text of the manuscript, we calculate JEV transmission suitability using predictions of *Culex* mosquito and ardeid bird habitat suitability relevant to February 2022 (Figure 4). We choose this set of predictions as they capture the most extreme conditions of the 2021–2022 summer and preceded the months when the majority of JEV detections were recorded during the outbreak; conditions during the period preceding the detections are assumed to have facilitated the wide geographical spread of the outbreak. However, it is possible to choose alternative maps of *Culex* mosquito and ardeid bird habitat suitability provided by models described in Duncan et al. [8]. In Supplementary Figure S5, we explore the use of median estimates of habitat suitability for the entire period of December 2021–March 2022. Medians are calculated from the set of monthly estimates produced by the Duncan et al. models. These surfaces are chosen as estimates of habitat suitability under average conditions for the entire summer, as distinct from the extreme conditions captured in the estimates in the main text. The scaled 2021–22 summer median map of *Culex* mosquito habitat suitability (Supplementary Figure S5(a)) is generally lower than the scaled February 2022 map in northern and central Australia, and higher in eastern Australia (Supplementary Figure S5(c)). Similarly, the 2021–22 summer median map of ardeid bird habitat suitability (Supplementary Figure S5(b)) is lower than the February 2022 map in northern and central Australia and higher in south-eastern Australia (Supplementary Figure S5(d)); the footprint of the Great Dividing Range is particularly clear here. Conditions were also more suitable under the December–March map in west and far north Western Australia and the Northern Territory near Darwin. In other words, conditions for both ardeid birds and *Culex* vectors were relatively better in northern Australia in the “extreme” February map, and relatively better in south-eastern Australia in the “average” December–March median map. This is interesting given the majority of initial JEV detections were recorded in south-eastern Australia (where surveillance effort, and subsequently power to detect transmission, are higher; Figure 5(b)).

We calculate JEV transmission suitability for a transmission cycle of ardeid birds and *Culex* vectors only using the December–March median maps (Supplementary Figure S5(e)), to complement the equivalent surface using the February-only maps (Figure 4(b)). JEV transmission suitability is estimated to be higher under February-only input maps, relative to December–March median maps, in northern Australia, particularly along the inland waterways highlighted in the ardeid bird habitat suitability map (Supplementary Figure S5(f)). JEV transmission suitability is estimated to be higher under December–March median maps, relative to February-only maps, in eastern Australia, including south-east Queensland, and in western and far north Western Australia.

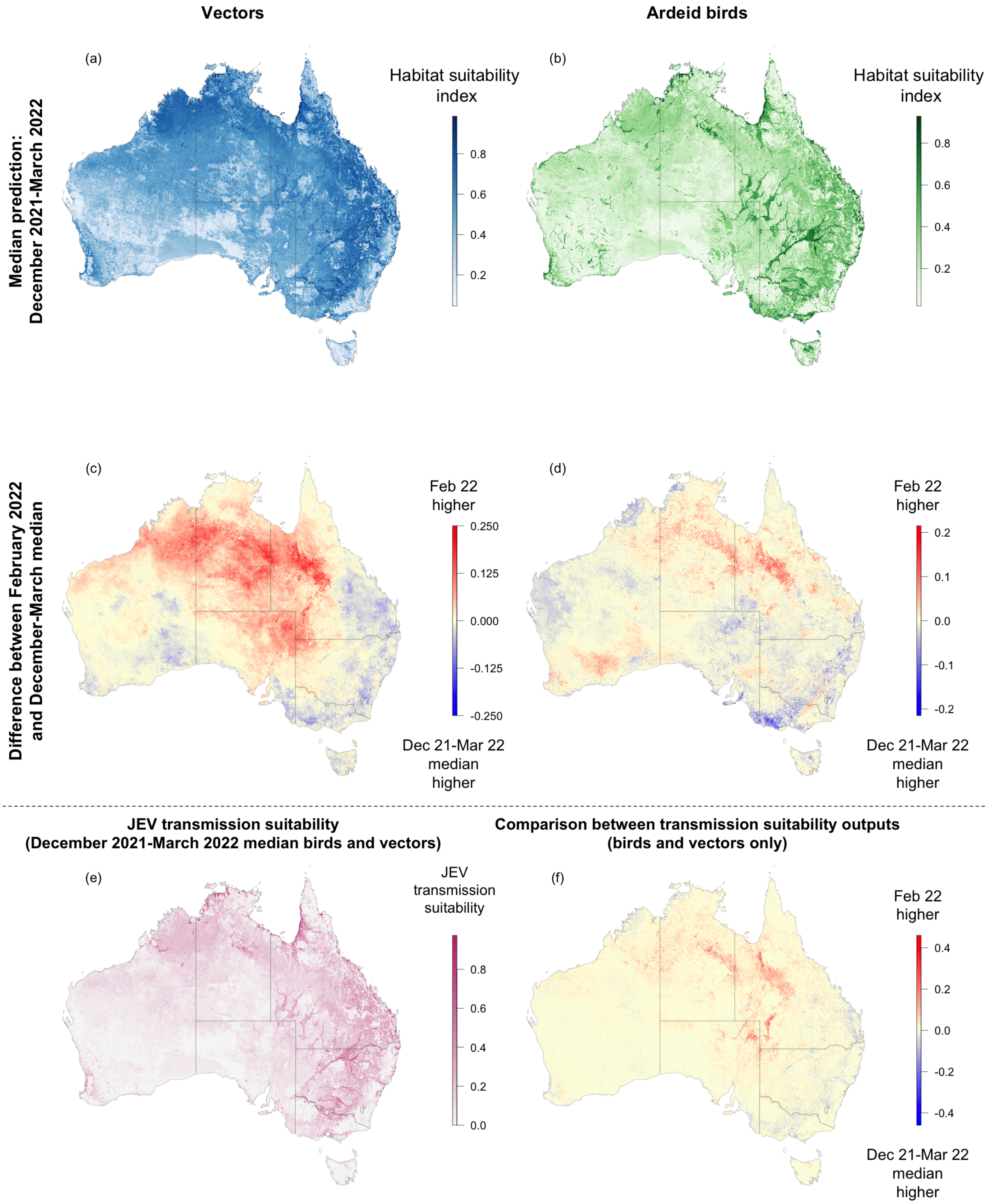

**Figure S5: Alternative estimate of JEV transmission suitability (joint habitat suitability for ardeid birds and *Culex* mosquitoes) using median estimated habitat suitability for birds and vectors from December 2021–March 2022.** Median predicted habitat suitability for (a) *Culex* mosquitoes and (b) ardeid birds from December 2021–March 2022, generated using models described in Duncan et al. [8]; difference between predicted habitat suitability for February 2022 (as employed in main manuscript) and for December 2021–March 2022 median for (c) *Culex* mosquitoes and (d) ardeid birds; (e) JEV transmission suitability using median estimated habitat suitability for birds and vectors from December 2021–March 2022; (f) difference between surface in (e) and equivalent surface using February 2022 predictions only (Figure 4(b)).

### S4 Notes on serological testing and diagnostic uncertainty

#### S4.1 Background

The dominant tools for diagnosing JEV infection in humans and other mammals in Australia are serological tests for JEV antigens or antibodies in blood or cerebrospinal fluid samples [9]. Serological investigations are recommended for use in clinical diagnosis and surveillance of JEV by the World Health Organization [10] and the World Organisation for Animal Health [11]. It is well recognised that diagnoses are complicated by cross-reactivity with antibodies of closely related flaviviruses (e.g., Murray Valley encephalitis virus (MVEV) and the Kunjin strain of West Nile virus ($\mathrm{WNV_{KUNV}}$)).

A variety of serological tests have been employed for JEV surveillance across Australian jurisdictions and host types, selected due to their differing test specificity and sensitivity, costs, sample types required, availability of contrast or control samples, laboratory expertise, and biosecurity clearance. Diagnostic algorithms for serological testing have been developed to support clinical investigations in humans, pigs, and other domestic animals (e.g.,[12]). With hosts that can be monitored in a clinical context, or that are captive/domesticated, cross-reactivity that may confound an isolated test result can be addressed by repeat sampling ('rebleeds') and testing to detect rising antibody responses such that JEV antibody titres increase dramatically (fourfold is the standard) while other flavivirus antibody titres do not. With a pathogen like JEV that only sporadically spills over from a wildlife reservoir, opportunities for repeat sampling of individual hosts are usually limited to sentinel animals and clinical investigations of domestic animals.

Where repeat sampling is not practical or affordable, cross-reactivity is addressed by serial testing of one serum sample, contingent on earlier test outcomes. An initial reactive pan-flavivirus or JEV assay might be followed by enzyme-linked immunosorbent assay (ELISA) tests targeted at candidate flavivirus antibodies conducted in parallel. In this context the fourfold threshold is typically applied in a static contrast, where a fourfold greater titre for JEV antibody than for MVEV or $\mathrm{WNV_{KUNV}}$ antibodies is considered indicative of JEV exposure. A fourfold contrast with high titre might suggest a relatively recent infection, whereas that magnitude difference with lower overall titres may suggest either an early stage IgG antibody response or historical infection.

Slightly different procedures might be available depending on the host type [12], and on the timing of the sample with respect to a suspected JEV infection. Tests that are available for sera sampled soon after infection use IgM antibodies that have relatively lower specificity [9]. Equivalent tests conducted on IgG antibodies are likely to be more reliable, but these antibodies rise later and consequently are less likely to be in samples obtained outside of the context of clinical investigation.

The preferred reference standard for serological testing is the plaque-reducing neutralisation test (PRNT) which tests the functional ability of antibodies in a sample to neutralise live virus particles [13, 14]. This test is the most specific, the most expensive [12], and can be offered only by facilities accredited to handle infectious JEV at biosecurity level/personal containment 3, such as the Australian Centre for Disease Preparedness (ACDP) and State Government reference laboratories. Not all samples in the data we received had been subject to tests of equivalent specificity and sensitivity, or the ideal PRNT confirmation. The unprecedented testing demand created by the JEV outbreak had exhausted laboratory testing capacity and consequently a number of datasets contain records that are unresolved pending PRNT analysis. In the absence of a JEV-positive or JEV-negative outcome, these records were excluded from our analysis.

#### S4.2 Semi-subjective weighting method

The model we use to infer transmission intensity accounts for differing rates of infection prevalence in different host types (see Methods). To incorporate serological data from non-human hosts into this model, we also account for the varying reliability of different diagnostic testing procedures. Cross-reactivity across JEV, MVE, and $\mathrm{WNV_{KUNV}}$ in serological assays is a well recognised issue, affecting the specificity of serological assays [13]. Testing protocols have been carefully developed to maximise test accuracy. However, specific estimates of sensitivity and specificity for each combination of species (i.e., domestic pigs, and chickens) and diagnostic type are lacking. In the absence of this information, we assign a semi-subjective "weight" to each type of diagnostic result (both positive and negative) reflecting the relative strength of the evidence provided by a sample of each type regarding the presence or absence of current or previous JEV infection in the host. Our proposed schema of weights for all combinations of non-human host types and diagnostic results is shown in Supplementary Table S1. Confirmed human cases are assigned a weight of one (divided across multiple exposure sites, where provided). We do not include inconclusive serological results in our model.

**Table S1: Schema of likelihood weights applied to different types of diagnostic testing procedure and non-human host species when incorporated in the transmission intensity models.** Confirmed human cases are also included, with a weight of one. Choice of weights is semi-subjective, with lower weights reflecting lower levels of diagnostic certainty. Weights can be interpreted in terms of the equivalent "number" of data points, i.e., two samples each with a weight of 0.5 have equivalent impact in the model as a single datapoint with weight one. OFV = other flavivirus (e.g., MVEV, $WNV_{KUNV}$). Greater than or equal to ($\geq$) indicates a greater than or equal to 4-fold difference in titre. Inconclusive refers to any results where there is not at least a 4-fold difference, including where titres are very high or very low for both JEV and OFV. Note that the collated dataset does not include all diagnostic types for all host types.

| | | | | ELISA-only | | | PRNT | | |
|---|---|---|---|---|---|---|---|---|---|
| | PCR +ve | PCR -ve | Initial sero. screen -ve | OFV $\geq$ JEV | JEV $\geq$ OFV | Inconcl. | OFV $\geq$ JEV | JEV $\geq$ OFV | Inconcl. |
| **Result** | Positive | Negative | Negative | Negative | Positive | Not used | Negative | Positive | Not used |
| **Weight** | | | | | | | | | |
| Human | 1 | - | 1 | 0.5 | 0.5 | - | 1 | 1 | - |
| Domestic pig | 1 | 1 | 0.5 | 0.25 | 0.25 | - | 0.5 | 0.5 | - |
| Feral pig | 1 | 1 | 0.5 | 0.25 | 0.25 | - | 0.5 | 0.5 | - |
| Chicken | 1 | 1 | 0.5 | 0.25 | 0.25 | - | 0.5 | 0.5 | - |
| Mosquito | 1 | 1 | - | - | - | - | - | - | - |

#### S4.3 Treatment of tests with conflicting results

In some cases, domestic or feral pigs were tested by both serological and molecular methods. Where any animal was tested by both serological and molecular methods and the outcomes of the test were in conflict, the positive result (or "exposure") was retained (Supplementary Figure S6), with its associated diagnostic weight (Table S1). For example, if an animal was tested by PCR, returning a negative result, and by PRNT, returning a positive result, the record was treated as a JEV exposure, with diagnostic weighting of 0.5. This treatment of the data is included in acknowledgement of the wider possible time period of exposure captured in a positive serological test: a negative test result by PCR need not indicate that an animal was not recently exposed to JEV [15].

The number of animals with conflicting results, where an animal with a negative PCR test is "promoted" by a positive serology-based test, is displayed in Table S2. The transmission intensity model, fitted to the dataset without the additional "promoted" seropositives, is displayed in Supplementary Figure S7. The overall shape of transmission intensity in Supplementary Figure S7 is comparable to Figure 5, with some small differences around the sites of omitted seropositives: Figure 5 shows relatively greater transmission intensity at the Victoria/New South Wales border and relatively greater transmission intensity in north-central Queensland.

#### S4.4 In summary

Our use of likelihood weights is a pragmatic and conservative approach to synthesising multiple types and sources of serological data in the absence of detailed information on rates of diagnostic sensitivity, specificity, and misclassification rates of pathogens under the different diagnostic assays, and on the multi-assay diagnostic cascades used in different jurisdictions (Supplementary Figure S6). Where reliable sensitivity and specificity data are available, more principled modelling approaches exist to explicitly account for these known testing issues and estimate sero-prevalence of a given pathogen [16]. In the case of JEV, where test specificity depends on both cross-reaction rates and the prevalences of other flaviviruses, reliable estimation of spatial variation in seroprevalence of JEV would require simultaneous estimation of the sero-prevalence of other flaviviruses. This simultaneous estimation is complicated by the fact that the various flaviviruses

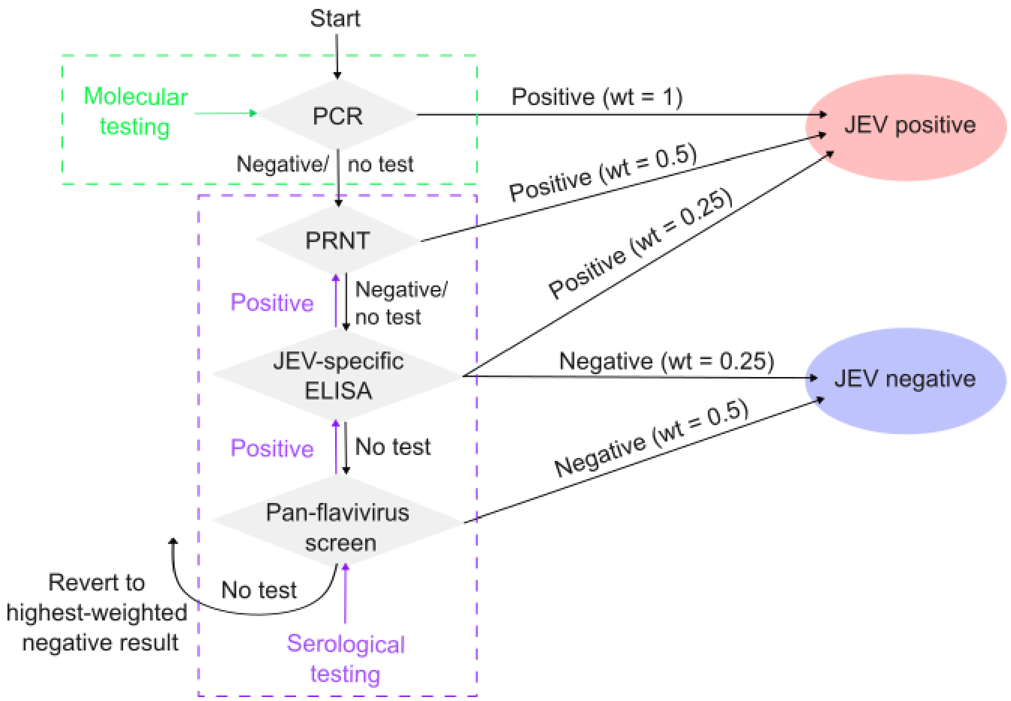

**Figure S6: Categorisation of domestic and feral pigs subjected to multiple forms of JEV surveillance.** By following the flow diagram indicated in black, each animal is assigned a test outcome of either "JEV positive" (indicating *either* detection of JEV or antibodies for JEV) or "JEV negative". Semi-subjective weights specific to the host type (feral or domestic pig), test outcome, and diagnostic method are indicated (Table S1). A positive result with associated lower diagnostic certainty is prioritised over a negative result with higher diagnostic certainty. Examples of "diagnostic cascades" in the data generation process are overlaid: molecular screening involved a single test (green), whereas under the serological screening pathway, JEV-specific ELISA and/or PRNT screening may have been conducted *only if* a sample was associated with a positive pan-flavivirus screening (purple).

| Host | NSW | Vic | Qld | SA | WA | Tas | NT |
|---|---|---|---|---|---|---|---|
| **Humans** - PCR and serological test data | | | | | | | |
| Potential exposure sites | 19 | 25 | 16 | 14 | 0 | 0 | 3 |
| **Mosquitoes** - PCR test data | | | | | | | |
| JEV negative | 2,336 | 4,196 | 5,974 | 1,254 | 1,024 | - | 747 |
| JEV PCR positive | 3 | 7 | 4 | 2 | 0 | - | 9 |
| **Domestic pigs** - PCR and serological test data | | | | | | | |
| JEV negative | 70 | 66 | 91 | 32 | 12 | 8 | 3 |
| JEV antibody positive | 27 | 6 | 6 | 4 | 0 | 0 | 0 |
| JEV PCR positive | 29 | 23 | 19 | 10 | 0 | 0 | 0 |
| **Sentinel chicken flocks** - Serological test data | | | | | | | |
| JEV negative | 187 | - | - | 279 | 273 | - | 25 |
| JEV antibody positive | 1 | - | - | 2 | 6 | - | 0 |
| **Feral pigs** - PCR and serological test data | | | | | | | |
| JEV negative | 110 | ~~2~~ 0 | 475 | ~~2~~ 1 | 125 | 0 | 649 |
| JEV antibody positive | 85 | ~~0~~ 2 | 8 | ~~2~~ 3 | ~~1~~ 2 | 0 | 41 |
| JEV PCR positive | 0 | 0 | 6 | 0 | 0 | 0 | 59 |

**Table S2: Summary of collated national JEV surveillance/detection data by host and jurisdiction over the period Jan 2021–Nov 2023, with alternative treatment of multiple tests with conflicting results.** Counter factual to Table 2: where two tests coincided using multiple screening methods with conflicting results, the positive result is prioritised, according to Supplementary Figure S6 (blue). Changes to the Table 2 when results with the highest diagnostic weight are prioritised (i.e., negatives by PCR over JEV antibody-positive tests) are indicated in red.

are likely to be spatially correlated (e.g. all are more prevalent in areas with higher mosquito abundances), but would be achievable with diagnostic testing procedures that report results for multiple flaviviruses. Whilst this was the case for some serological assays, most diagnostic cascades relied on an initial serological screen for JEV, which limits the utility of the data for disentangling the distribution of JEV from the other flaviviruses. In the absence of such data on test sensitivity and specificity for JEV with co-circulating flaviviruses, alternative sets of weightings could have been designed to represent different ideas about test sensitivity and specificity for JEV, or serological test interpretation more broadly given geographic context or repeat sampling in the case of sentinel animals. The sensitivity of the model output to those different sets of assumptions could have been explored and quantified, however such treatment was beyond the scope of this work.

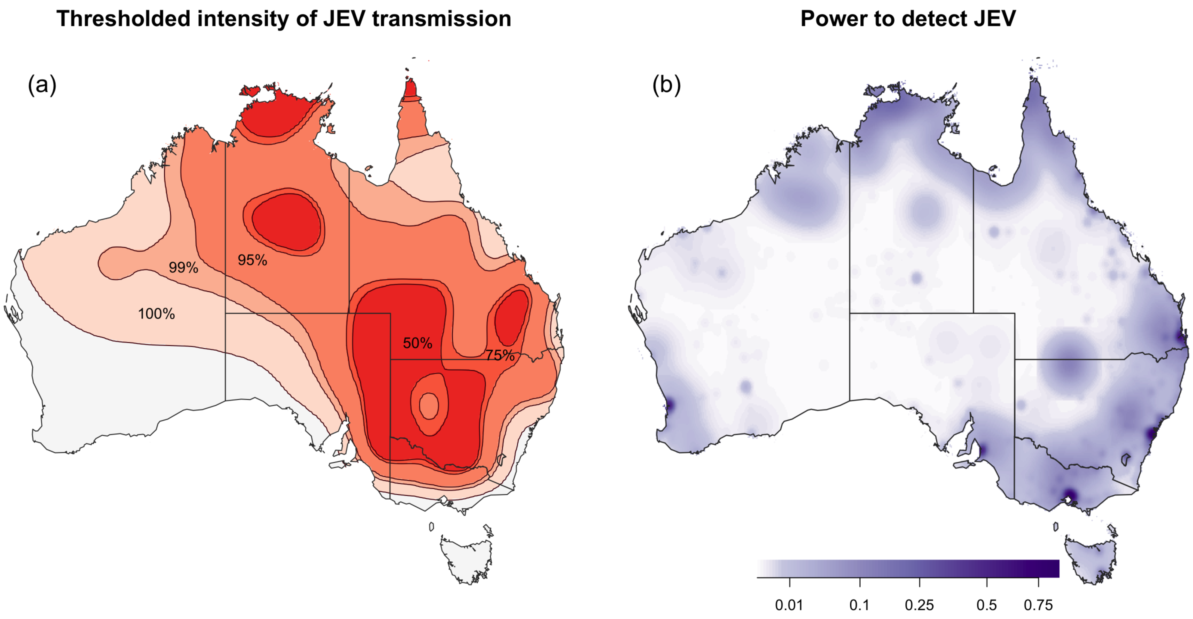

**Figure S7: Estimated transmission intensity and power to detect JEV under alternative treatment of simultaneous tests with conflicting results.** Counterfactual to Figure 5: where an animal is tested simultaneously by molecular and serological assay, the result of the molecular assay is prioritised without regard to the result of the serological assay.

### S5 Detailed description of power to detect JEV transmission

We include a detailed description of the calculation of power to detect JEV transmission during the outbreak below. Using data of locations of surveillance effort, by detection group and diagnostic type, we produced maps of statistical power to detect JEV transmission in each detection group and across all JEV surveillance, regardless of detection group. These maps can be interpreted as the probability that a JEV detection of any type would occur in a given pixel, if JEV transmission were present at relatively high intensity.

Recall, in Eq (2), we assumed that the number of JEV detections at a given location, $i$, for a given detection group (of humans, mosquitoes, domestic pigs, feral pigs, or sentinel chickens), $g$, for a given diagnostic type (either PCR, serology, or unknown test type), $d$, follows a Poisson distribution, with a rate equal to the expected number of JEV detections for a given location, group, and diagnostic method. We find the median of the estimated transmission intensity surface at the set of locations associated with a JEV detection, $\lambda_{50}$:

$$\lambda_{50} = \text{med}(\{\exp(\hat{f}(\boldsymbol{x}_i)) \ \forall \ i \text{ coinciding with JEV detection}\}). \tag{S1}$$

This median value is the level of estimated transmission intensity corresponding to the 50% threshold extent. To estimate the expected number of JEV detections, assuming JEV is present, we substitute the median estimated transmission suitability in the presence of a detection, $\lambda_{50}$, into transmission intensity, $f(x)$, in Eq (2).

The distribution of JEV detections, given JEV is present, is then:

$$\hat{y}_{i,g,d} | \lambda_{50} \sim \text{Pois}\Big( \exp\big(\log(\lambda_{50}) + \beta_g + \gamma_d\big) \phi_{i,g} \Big). \tag{S2}$$

Note that as $\lambda_{50}$ is constant for all $i$, the only source of spatial variation in Eq (S2) and resulting power-to-detect outputs is spatial variation in surveillance effort, $\phi_{i,g}$. The power to detect JEV for each location and detection group is the complement of the probability of no detection for any diagnostic type, given JEV is present:

$$\begin{aligned} \Pr(\hat{y}_{i,g} > 0 | \lambda_{50}) &= 1 - \Pr(\hat{y}_{i,g} = 0 | \lambda_{50}). \\ &= 1 - \prod_{d \in D_g} \Pr(\hat{y}_{i,g,d} = 0 | \lambda_{50}), \end{aligned} \tag{S3}$$

where $D_g$ is the set of relevant diagnostic types for the detection group (e.g. there is no serology-based surveillance in mosquitoes in our dataset). Overall statistical power to detect JEV, across each of the four modelled detection groups and three diagnostic methods then follows, assuming the probability of detecting JEV in one group is independent of detection in others:

$$\begin{aligned} \Pr(\hat{y}_i > 0 | \lambda_{50}) = 1 - &\Pr(\hat{y}_{i,\text{human}} = 0 | \lambda_{50}) \times \Pr(\hat{y}_{i,\text{mosquito}} = 0 | \lambda_{50}) \\ &\times \Pr(\hat{y}_{i,\text{domestic pig}} = 0 | \lambda_{50}) \times \Pr(\hat{y}_{i,\text{feral pig}} = 0 | \lambda_{50}) \\ &\times \Pr(\hat{y}_{i,\text{chicken}} = 0 | \lambda_{50}). \end{aligned} \tag{S4}$$

We estimate species group-level power to detect transmission in a nominal 11 × 11 km$^2$ pixel, given transmission intensity is relatively high (i.e., $\Pr(\hat{y}_{i,g} > 0 | \lambda_{50})$) in Supplementary Figures S8. The power to detect transmission is greatest in feral pigs (Supplementary Figure S8(d)) and humans (Supplementary Figure S8(a)): in humans, we have assumed passive surveillance effort proportional to human population density, so that high detection probabilities are concentrated in capital cities and are relatively low elsewhere. Outside of a capital city, the probability of detecting JEV in humans, given surveillance intensity is proportional to human population density *and* given that JEV transmission is at high intensity is less than 0.001 positives per "test". In feral pigs, high estimated power-to-detect is reflective of positive detections of JEV in areas with relatively low surveillance effort (at least as surveillance effort is captured in the dataset; Figures 2(d) and 3(d), and Supplementary Figure S10(d)), in South Australia and Victoria (Table 2).

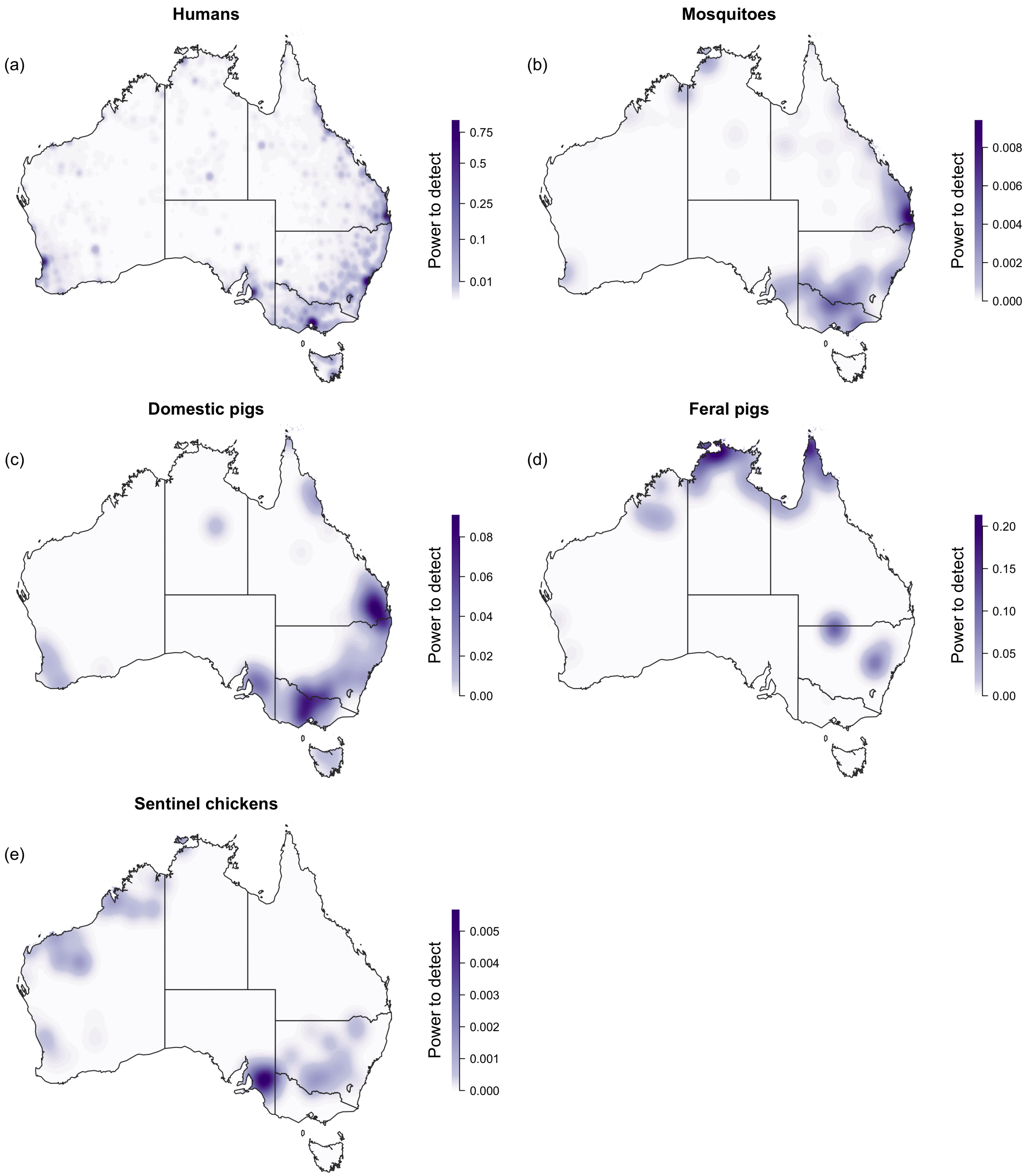

**Figure S8: Estimated species group-level power to detect transmission.** Probability of detecting JEV in (a) humans, (b) mosquitoes, (c) domestic pigs, (d) feral pigs, and (e) sentinel chickens, given recorded surveillance effort and given that JEV transmission intensity is equal to $\lambda_{50}$, the median estimated transmission intensity across all locations where a JEV detection was recorded.

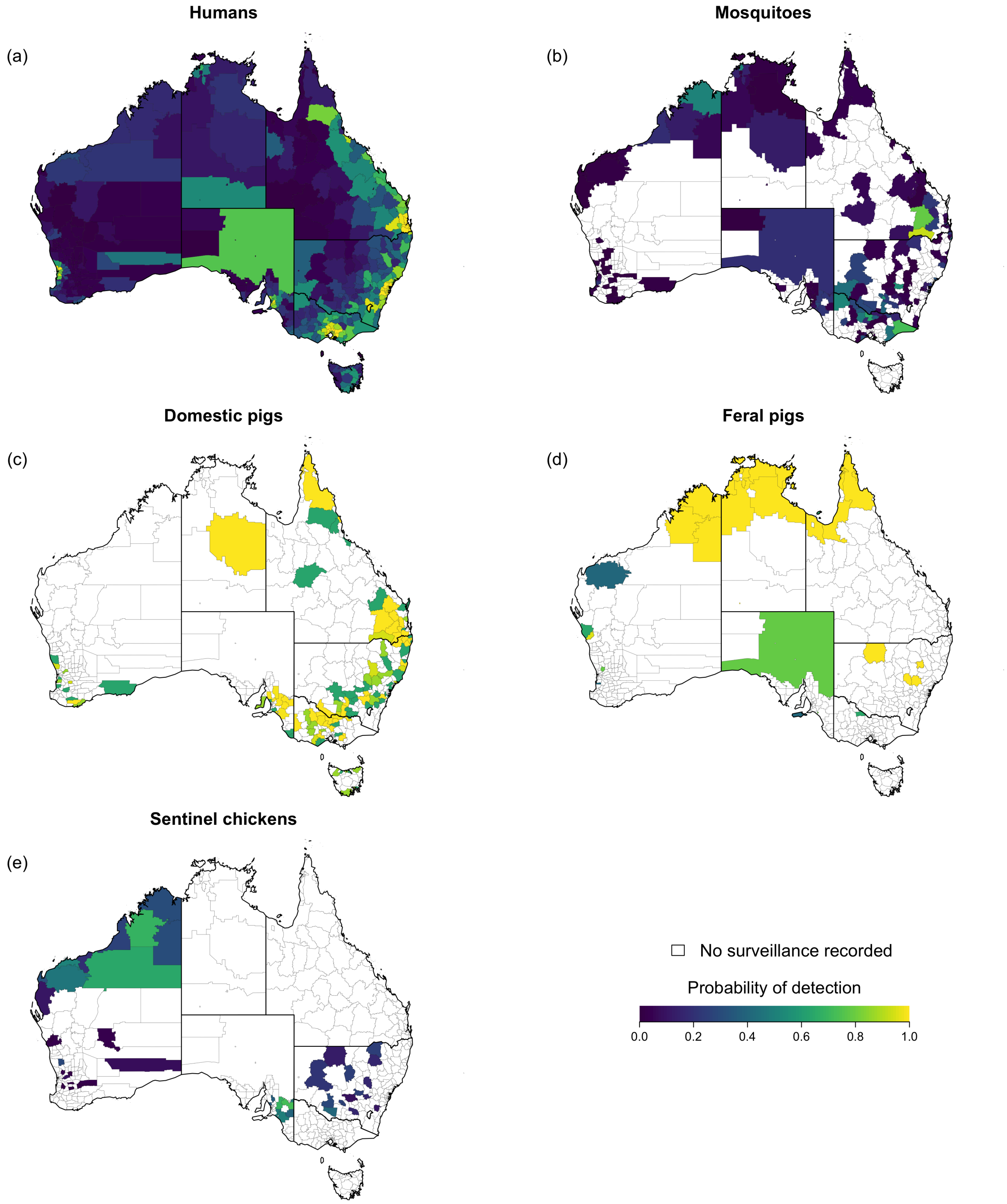

**Figure S9: Estimated species group-level power to detect transmission in Australian local government areas.** Probability of detecting JEV in (a) humans, (b) mosquitoes, (c) domestic pigs, (d) feral pigs, and (e) sentinel chickens during the 2021–2023 outbreak, in Australian local government areas given recorded surveillance effort and given that JEV transmission intensity is equal to $\lambda_{50}$, the median estimated transmission intensity across all locations where a JEV detection was recorded. Note that estimates are based on surveillance activities during the 2021–2023 outbreak, which may not reflect current surveillance programs. For example, post-2023 some jurisdictions have replaced sentinel chicken surveillance in favour of other surveillance methods (e.g. molecular testing on trapped mosquitoes).

### S6 Further supplementary figures and tables

**Table S3: Spatial predictor variables contributing to national models of ardeid and *Culex* habitats used in this work.** Reproduced from Duncan et al. [8].

| Variable | Definition | Source |
|---|---|---|
| 'Static' fractional land cover (0–1) | | |
| Frequency of indundation | Base 30 m product represents for each pixel the proportion of all cloud-free images in which it was deemed occupied by water for the period 1987–2021. | GeoSciences Australia 'Water Observations from Space' [17]. The original product was summarised to average frequency calculated for 1 km grid. |
| Permanent water | Area covered for ≥9 months of year by fresh or salt water. | European Space Agency [18]. The original 10 m categorical product (see Table 2 of [19]) was aggregated to proportional cover of each type 1 km grid. |
| Herbaceous wetland | Natural herbaceous vegetation permanently or regularly flooded. | |
| Mangrove | Intertidal zones dominated by treed vegetation. | |
| Grassland | Area dominated by herbaceous vegetation of ≥ 10% cover, may include uncultivated pasture or crop. | |
| Cropland | Occupied by annual crops harvested at least once within 12 months of sowing. | |
| Bare / sparse | Exposed substrate, always less than 10% vegetated cover. | |
| Shrubland | Natural shrub cover of ≥ 10% or more. | |
| Forest | Tree cover of ≥ 10% cover. Plantations, swamp-forest captured in this class and canopy may cover other types (e.g., water bodies). | |
| Temporally dynamic environmental variables | | |
| Night temperature (Night T °C) | Average Land-surface temperatures derived from MODIS v6 8-daily 1 km product MOD11A2 | MODIS base products were post-processed by a global implementation of Weiss et al [20] algorithm to gap-fill cloud-affected scenes. The gap-filled outputs were aggregated to a monthly mean. |
| Day temperature (Day T °C) | | |
| EVI (Greenness) | Enhanced Vegetation Index - composite monthly product robustly emphasising vegetation canopy density and greenness against soil and structure background variation (see MOD13A2 [21]) | Regenerated from raw MODIS products MCD43D62–MCD43D68 and post-processed by a global implementation of Weiss et al [20] algorithm to gap-fill scenes affected by cloud. The gap-filled outputs were aggregated temporally to a monthly mean |
| TCW (Wetness) | Tasselled Cap Wetness - Reductive transformation and rotation of Landsat TM reflectance bands emphasising moisture variation in soils and possibly vegetation [22] | |
| TCB (Brightness) | Tasselled Cap Brightness - Weighted sum of Landsat TM reflectance bands emphasising variation in soil physical properties [22] | |
| Rainfall (mm) | Monthly rainfall total (mm) according to the interpolation algorithm of Climate Hazards group Infrared Precipitation with Stations (CHIRPS) [23]. | Resampled to 1 km grid. |
| Rainfall (lag 1 month) | Monthly rainfall total (mm) as above from CHIRPS for the month preceding the observation. | |

**Table S4: Summary of virus surveillance data contributing to the manuscript, including systematic survey and routine surveillance for vector-borne diseases including JEV, and detections of JEV for the period December 2021–October 2023.** Truncated lats/longs are longitude and latitude coordinates to one decimal place (an area of approximately 11 km × 11 km). NAQS = Northern Australia Quarantine Strategy. LGA = Local Government Area.

| Type & Region | Source | Spatial precision | Temporal precision |
|---|---|---|---|
| Flavivirus detections in humans | | | |
| National | National Notifiable Diseases Surveillance System (NNDSS) [24] | postcode | date |
| NSW | NSW Health | townships | date |
| NT | NT Health | townships | date |
| QLD | Queensland Health | truncated lats/longs | date |
| SA | SA Health | lats/longs | month |
| VIC | Dept of Health | truncated lats/longs | date |
| *There were no detections in ACT, TAS, or WA throughout 2021–2023.* | | | |
| Commercial pig premises | | | |
| Notifiable detections of JEV | | | |
| NSW | Dept of Primary Industries & Regional Development | truncated lats/longs | date |
| QLD | Dept of Primary Industries | truncated lats/longs | date |
| SA | Dept of Primary Industries & Regions SA | LGA-level | date |
| VIC | Agriculture Victoria | truncated lats/longs | date |
| *There were no notifiable detections in NT, TAS, WA pig premises throughout 2021–2023.* | | | |
| JEV-delimiting serological survey | | | |
| NSW | Dept of Primary Industries & Regional Development | LGA | lab test date |
| QLD | Dept of Primary Industries | LGA | lab test date |
| SA | Dept of Primary Industries & Regions SA | LGA | lab test date |
| VIC | Agriculture Victoria | LGA | lab test date |
| WA | Dept of Primary Industries & Regional Development | LGA | lab test date |
| TAS | Animal Health Aust. / Dept of Agriculture, Fisheries & Forestry | LGA | lab test date |
| Arbovirus surveillance: Sentinel chicken flocks | | | |
| NSW | NSW Health (online) [25] | township | date |
| NT | NAQS | truncated lats/longs | date |
| SA | Dept of Health & Wellbeing | postal area | date |
| WA | Dept of Health | truncated lats/longs | date |
| *Flocks not maintained in ACT, QLD, TAS, and VIC* | | | |
| Other testing of domestic, feral or wild animals | | | |
| QLD, NT, WA | NAQS | lats/longs | date |
| NSW | Dept of Primary Industries & Regional Development | truncated lats/longs | date |
| SA | Dept of Primary Industries & Regions SA | lats/longs | date |
| VIC | Agriculture Victoria | truncated lats/longs | date |
| Arbovirus surveillance: Mosquitoes | | | |
| NSW | NSW Health | lats/longs | date |
| NT | NT Health | lats/longs | date |
| QLD | Queensland Health | truncated lats/longs | date |
| SA | SA Health | lats/longs | date |
| VIC | Dept of Health | lats/longs | date |
| WA | WA Health | lats/longs | date |
| *TAS pilot surveillance samples were not tested; ACT initiated arbovirus surveillance in 2022/23 but test result data were not obtained for this study.* | | | |

**Figure S10: Locations of JEV detections (red squares) and negative tests (blue squares).** Point opacity is determined by the diagnostic method associated with the surveillance record (Table S1) and the likelihood of JEV exposure at the location, where it was provided, or exposure weight. (a) Locations of probable exposure sites for confirmed cases of JEV in humans. (b) Locations and results of tests for JEV in mosquitoes. (c) Locations and results of tests for JEV in domestic pigs. (d) Locations and results of tests for JEV in feral pigs. (e) Locations and results of tests for JEV in sentinel chickens. (f) Distribution of national JEV surveillance through time, by host type. Locations of JEV detections only are visualised in Figure 2.

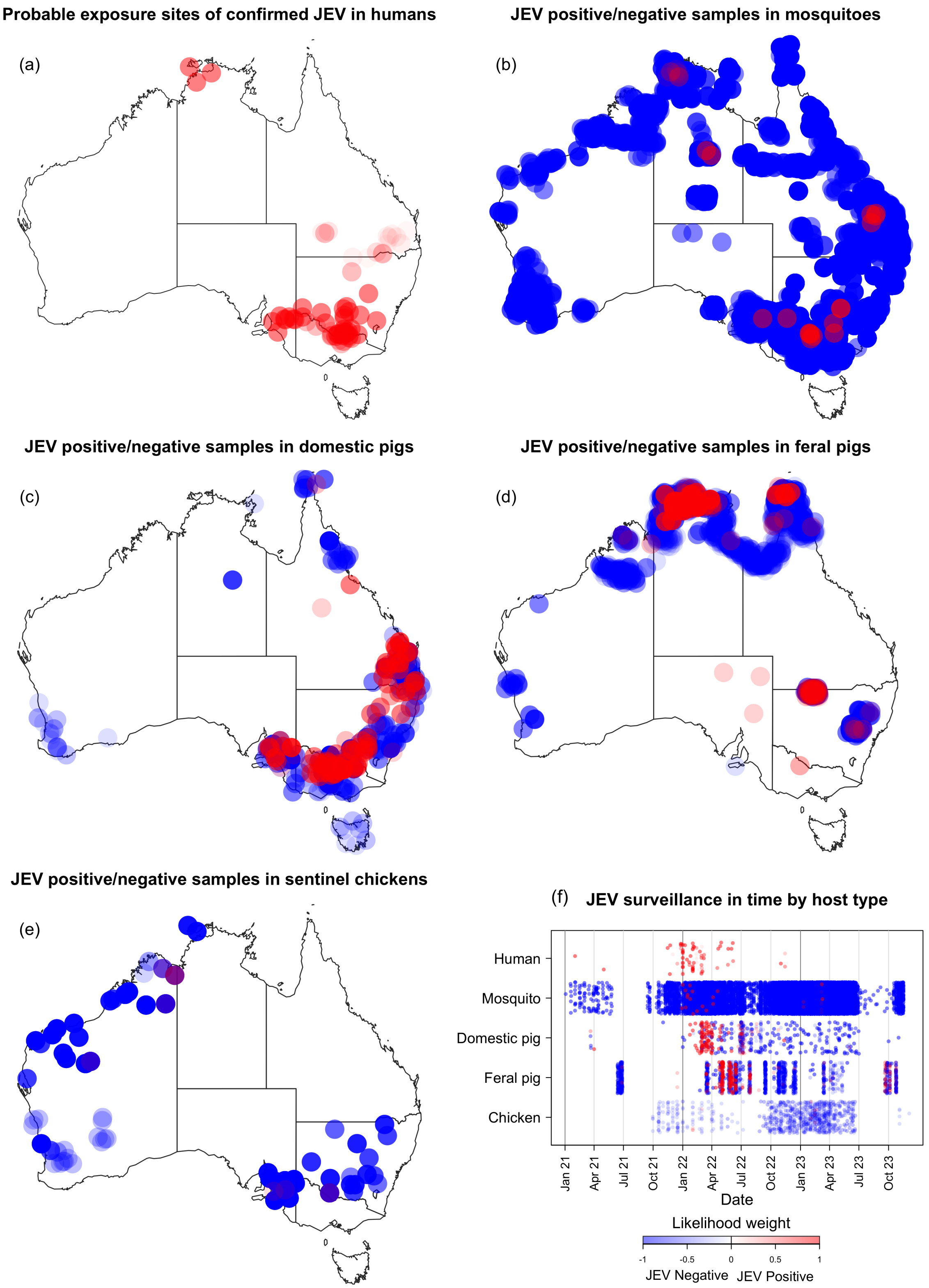
